# AI-Powered Segmentation and Prognosis with Missing MRI in Pediatric Brain Tumors

**DOI:** 10.1101/2025.07.14.25331187

**Authors:** Dimosthenis Chrysochoou, Deep B. Gandhi, Sahand Adib, Ariana M. Familiar, Neda Khalili, Nastaran Khalili, Jeffrey B. Ware, Wenxin Tu, Paarth Jain, Hannah Anderson, Shuvanjan Haldar, Phillip B. Storm, Andrea Franson, Michael Prados, Cassie Kline, Sabine Mueller, Adam Resnick, Arastoo Vossough, Christos Davatzikos, Ali Nabavizadeh, Anahita Fathi Kazerooni

**Affiliations:** University of Pennsylvania, Department of Bioengineering, Philadelphia, PA, USA; Center for Data-Driven Discovery in Biomedicine (D3b), Children’s Hospital of Philadelphia, Philadelphia, PA, USA; Department of Neurosurgery, Children’s Hospital of Philadelphia, Philadelphia, PA, USA; Department of Radiology, Perelman School of Medicine, University of Pennsylvania, Philadelphia, PA, USA; Department of Pediatrics, University of Michigan Medical School, Ann Arbor, MI, USA; Division of Radiology, Children’s Hospital of Philadelphia, Philadelphia, PA, USA; Division of Oncology, Children’s Hospital of Philadelphia, Philadelphia, PA, USA; Department of Neurological Surgery and Department of Pediatrics, University of California San Francisco, San Francisco, CA, USA; Department of Neurology and Pediatrics, University of California San Francisco, San Francisco, CA, USA; AI^2^D Center for AI and Data Science for Integrated Diagnostics, University of Pennsylvania, Philadelphia, PA, USA; Department of Neurosurgery, Perelman School of Medicine, University of Pennsylvania, Philadelphia, PA, USA

## Abstract

**Importance:** Brain MRI is the main imaging modality for pediatric brain tumors (PBTs); however, incomplete MRI exams are common in pediatric neuro-oncology settings and pose a barrier to the development and application of deep learning (DL) models, such as tumor segmentation and prognostic risk estimation.

**Objective:** To evaluate DL-based strategies (image-dropout training and generative image synthesis) and heuristic imputation approaches for handling missing MRI sequences in PBT imaging from clinical acquisition protocols, and to determine their impact on segmentation accuracy and prognostic risk estimation.

**Design:** This cohort study included 715 patients from the Children’s Brain Tumor Network (CBTN) and BraTS-PEDs, and 43 patients with longitudinal MRI (157 timepoints) from PNOC003/007 clinical trials. We developed a dropout-trained nnU-Net tumor segmentation model that randomly omitted FLAIR and/or T1w (no contrast) sequences during training to simulate missing inputs. We compared this against three imputation approaches: a generative model for image synthesis, copy-substitution heuristics, and zeroed missing inputs. Model-generated tumor volumes from each segmentation method were compared and evaluated against ground truth (expert manual segmentations) and incorporated into time-varying Cox regression models for survival analysis.

**Setting:** Multi-institutional PBT datasets and longitudinal clinical trial cohorts.

**Participants:** All patients had multi-parametric MRI and expert manual segmentations. The PNOC cohort had a median of three imaging timepoints and associated clinical data.

**Main Outcomes and Measures:** Segmentation accuracy (Dice scores), image quality metrics for synthesized scans (SSIM, PSNR, MSE), and survival discrimination (C-index, hazard ratios).

**Results:** The dropout model achieved robust segmentation under missing MRI, with ≤0.04 Dice drop and a stable C-index of 0.65 compared to complete-input performance. DL-based MRI synthesis achieved high image quality (SSIM > 0.90) and removed artifacts, benefiting visual interpretability. Performance was consistent across cohorts and missing data scenarios.

**Conclusion and Relevance:** Modality-dropout training yields robust segmentation and risk-stratification on incomplete pediatric MRI without the computational and clinical complexity of synthesis approaches. Image synthesis, though less effective for these tasks, provides complementary benefits for artifact removal and qualitative assessment of missing or corrupted MRI scans. Together, these approaches can facilitate broader deployment of AI tools in real-world pediatric neuro-oncology settings.

## INTRODUCTION

Multiparametric MRI plays a critical role in the evaluation of pediatric brain tumors (PBTs), supporting comprehensive assessment for diagnosis, treatment planning, radiomic analysis, and response monitoring^1^. Specifically, accurate response assessment depends on precise delineation of intratumoral subregions, including contrast-enhancing and non-enhancing tumor, cystic components, and peritumoral edema, because each must be selectively included or excluded according to Response Assessment in Pediatric Neuro-Oncology (RAPNO) guidelines across multiple PBT histologies^2–7^.However, in clinical practice, complete MRI acquisitions are frequently unavailable, due to protocol variations or imaging artifacts. This poses a significant challenge for deep learning (DL) segmentation models trained on fully sampled, curated datasets ^8–11^. Protocols designed for clinical tasks such as surgical navigation often acquire only a limited subset of sequences, such as T1-weighted post-contrast and T2-weighted images. This is further exacerbated in pediatrics, where inherently small cohort sizes, due to the lower incidence of PBTs ^12^, are further reduced by excluding patients with missing sequences, restricting sample size and limiting model generalizability.

In recent years, a growing number of studies have addressed missing MRI sequences using generative DL models that synthesize missing scans from available inputs. These include architectural approaches, based on Generative Adversarial Networks (GANs)^13–15^, Denoising Diffusion Probabilistic Models (DDPMs)^16–18^, and transformer-based frameworks^19^, among others ^20–22^. An alternative class of methods includes segmentation models inherently robust to missing sequences, eliminating the need for synthesis. Strategies include disentangling modality-specific and shared features^23^, learning shared representations less dependent on any single input^24^,^25^ adversarial training to help networks learn effectively from both complete and incomplete MRI data^26^, style transfer techniques to bridge gaps between available and missing modalities^27^, and region-specific fusion methods guided by anatomical information^28^. However, many of these models are complex and computationally demanding, limiting their practical utility. A simpler and more effective inherently robust strategy was proposed in^29^ employing a dropout-like training mechanism that randomly removes input sequences during training to simulate missing data. Although effective, this model is not publicly available, hindering wider clinical adoption.

Most of these strategies have been developed and validated using adult cohorts, particularly the Brain Tumor Segmentation Challenge (BraTS) dataset ^30^ which is highly curated and may not adequately reflect the complexities encountered in real-world pediatric clinical practice. Moreover, no systemic comparison of generative models versus robustness-based approaches has been performed in pediatric tumor segmentation.

In this study, we address these gaps, using a multi-institutional, multi-histology PBT cohort to adapt a state-of-the-art generative model for MRI synthesis and to develop a dropout-trained automated segmentation pipeline, which we release publicly. To evaluate real-world clinical utility, we systematically assessed their performance in tumor segmentation across realistic missing MRI scenarios using retrospective clinical trial data from the PNOC003 and PNOC007 cohorts. Tumor volumes derived from these segmentations are subsequently integrated with electronic health record (EHR) data to facilitate downstream risk stratification.

## METHODS

### Datasets

We assembled a multi-institutional, multi-histology retrospective cohort of **715** pediatric patients from the Children’s Brain Tumor Network (CBTN)^31^,^32^ and the Brain Tumor Segmentation in Pediatrics (BraTS-PEDs)^8^ datasets. Additionally, we curated a retrospective longitudinal cohort of 43 patients with diffuse midline glioma (DMG), including diffuse intrinsic pontine glioma (DIPG), from the PNOC003 and PNOC007 clinical trials^33,34^ comprising 157 imaging timepoints. Each patient had standard MRI including pre- and post-contrast T1-weighted (T1w-pre, T1w-post), T2-weighted (T2w), and fluid-attenuated inversion recovery (FLAIR) images and expert manual segmentations. Demographics and preprocessing details are provided in eData, eTables 1,2.

### Automated Response Assessment Pipeline for Pediatric Brain Tumors

We developed an automated pipeline to assess treatment response in PBTs using standard MRI sequences (T1w-pre, T1w-post, T2w, and FLAIR) (Figure 1A). A DL segmentation model delineates key tumor subregions, including whole tumor (WT), enhancing (ET), and non-enhancing tumors (NET). From these segmentation masks, longitudinal tumor volumes, excluding edema, are computed at each timepoint. Volumetric features, combined with Electronic Health Record (EHR) data (age, sex, and treatment group), are input into a time-varying Cox regression model to estimate survival risk. Each component of the pipeline is detailed below.

**Figure 1.**
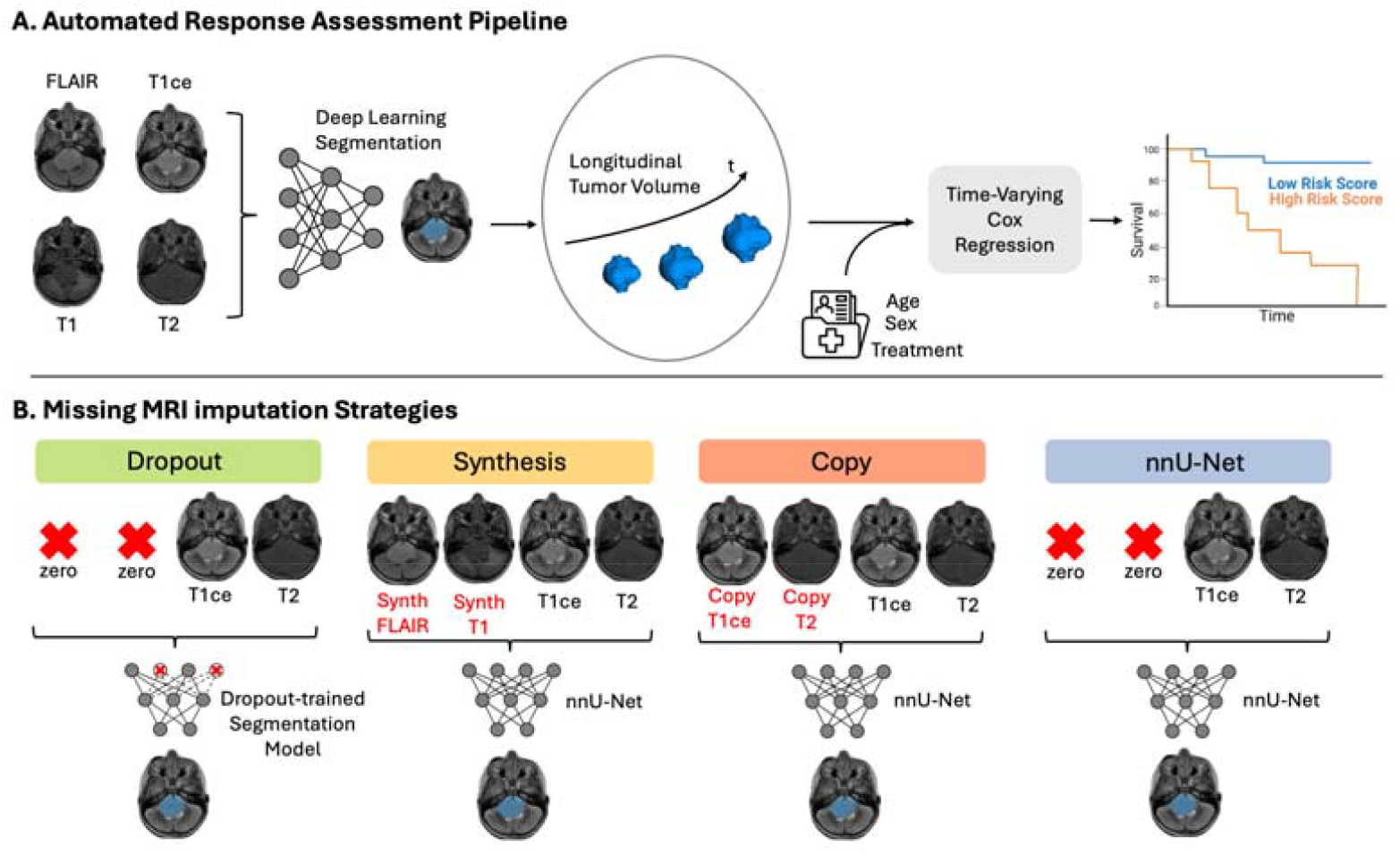
Overview of the Automated Response Assessment Pipeline: (A) Multiparametric MRI scans are segmented at each available timepoint using a deep learning model to extract longitudinal tumor volumes. Combined with electronic health record data, they are used in a time-varying Cox model to estimate the patient’s survival risk. (B) Strategies for handling missing MRI sequences during segmentation: *Dropout* uses a custom segmentation model trained with randomly dropped sequences enabling inherent robustness to missing inputs. *Synthesis, Copy*, and *nnU-Net* use the standard nnU-Net model downstream and impute missing sequences with synthetic scans, copies of closest physical counterparts, or zeros, respectively.

#### A. Deep Learning-Based Tumor Segmentation

We used the nn-UNet architecture ^35^ an open-source, self-configuring framework that automates all stages of medical image segmentation and has demonstrated state-of-the-art performance in adult and pediatric brain tumor segmentation^35,11^.

#### B. Time Varying Cox-Regression

To model survival, we implemented a time-varying Cox regression framework, appropriate for longitudinal clinical data where tumor burden may evolve over time. Following emerging evidence that volumetric measures better capture tumor growth and response than traditional bidimensional measurements^36,37^, tumor volume served as a time-varying covariate, while age, sex, and treatment group modeled as fixed effects.

### Handling Missing MRI Sequences in the Segmentation Pipeline

The pipeline (Figure 1A) uses four standard MRI sequences, but incomplete imaging is common due to protocols variability, artifacts, or in surgical navigation protocols. To address frequent absence of T1w-pre and/or FLAIR, we evaluated four strategies (Figure 1B): modality dropout, image synthesis, copy substitution, and zero-filled inputs with standard nnU-Net. Models were trained on 340 CBTN patients and fine-tuned on a validation set of 85 CBTN patients.

#### A. Modality Dropout: Training for Robustness to Missing Inputs

Inspired by the dropout mechanism used in neural networks to prevent overfitting ^38^, our “modality dropout” strategy randomly removes MRI sequences during training to simulate incomplete inputs, encouraging the model to learn representations robust to missing data.

We integrated this into the nnU-Net framework as a data augmentation step, preserving its automated configuration. For each training sample (comprising four MRI sequences), FLAIR and T1w-pre sequences are independently set to zero with probability p ∈ {0, 0.1, 0.2, …, 1}, treated as a tunable hyperparameter. For example, with *p* = 0.5 each sequence is dropped in 50% of samples, and both are dropped simultaneously in about 25% of cases, assuming independence. Due to the stochastic nature of this approach, the specific dropout patterns for a given patient vary across epochs, providing a diverse range of input combinations, helping the model generalize to real-world missing data scenarios. We optimized p by averaging WT Dice across four validation scenarios: (1) all sequences available, (2) FLAIR missing, (3) T1w-pre missing, and (4) both missing. The model trained with p=0.4 achieved the best average performance and was selected for inference on the test set.

#### B. Image Synthesis: Generating Missing Sequences with ResViT

As an alternative, we used generative model to synthesize missing sequences prior to segmentation. We selected ResViT^14^, a GAN-based publicly available model that has demonstrated strong performance in MRI synthesis on the adult BraTS dataset^30^. ResViT combines the sensitivity to global context of vision transformers, the local feature extraction capabilities of convolutional networks, and the image synthesis realism of adversarial learning strategies. We trained two ResViT models to respectively synthesize missing FLAIR from T2w and T1w-pre from T1w-post scans (Synth FLAIR and Synth T1, respectively, in Figure 1 B). These synthetic scans replaced missing sequences at nnU-Net inference. Following the tuning protocol described in ^14^, we performed a grid search over learning rates and pixel-wise loss weights, identifying optimal hyperparameters of 1e-4 and 200 respectively. Notably, these hyperparameters differ from those termed optimal in adult cohorts^14^.

#### C. Copy Substitution: A Simple Baseline Imputation

As a baseline, missing sequences were replaced with their closest physical counterparts^13^: missing FLAIR with T2w, and missing T1w-pre with T1w-post. This provides a benchmark to assess the added value of learned synthesis models, which are trained on the same input-output pairs. This comparison is critical, as DL-based synthesis models could converge to suboptimal solutions by simply replicating anatomical structures from the input without recovering the intended contrast, especially when source and target sequences are structurally similar. Superior performance by ResViT would suggest it captures contrast-specific features not present in the nearest physical modality.

#### D. Zeroed Input: Standard nnU-Net Without Imputation

As a final baseline, we evaluate the standard nnU-Net by running inference with missing sequences set to zero. This reflects the model’s default behavior without any imputation or robustness strategy and helps quantify the value added by the proposed imputation strategies.

Additional training details for all models are provided in eTraining Details, eFigures 1-3.

#### E. Code availability

The code for the Modality Dropout segmentation model will become publicly available upon publication of the paper. The remaining methods are publicly available from ^35^,^14^.

### Evaluation Experiments

To assess segmentation quality and clinical utility across models and imputation strategies, we conducted the following evaluations:

#### 1. Segmentation Performance

Dice scores for WT, ET, NET were calculated on held-out CBTN test data and the PNOC cohort. Sensitivity analysis and expert radiologist review focused on cases with the largest performance drop when T1w-pre or FLAIR were missing. Statistical significance was assessed using the Wilcoxon signed-rank test with Bonferroni corrections.

#### 2. Volumetric Accuracy and Survival Modeling (PNOC Clinical Data)

Using each segmentation approach of Figure 1B (with complete sequences or missing FLAIR) and expert manual segmentations, tumor volumes were computed and used to fit time-varying Cox models. Survival curves (Kaplan–Meier), forest plots, and hazard ratios were used to assess risk stratification performance. Kaplan–Meier curves report the concordance index (C-index) and log-rank p-value, reflecting model discrimination and survival separation, respectively. Patients were dichotomized into high/low-risk groups based on their predicted risk scores, with high-risk defined as risk ≥ median. Forest plots show hazard ratios with 95% confidence intervals (CI) and indicators of statistical significance per covariate. Differences in risk scores across Cox models were tested using the Wilcoxon signed-rank test with Bonferroni corrections (p=0.05).

3. Perceptual Quality Analysis of Synthesized scans

Image synthesis quality was evaluated using structural similarity index (SSIM), mean squared error (MSE), and peak signal-to-noise ratio (PSNR) on both CBTN and PNOC datasets.

## RESULTS

### Segmentation Performance

Figure 2 shows box-and-whisker plots of Dice scores for WT, ET, and NET across imputation strategies and missing MRI scenarios in the CBTN test set and PNOC clinical trial data. Median Dice scores are annotated, additional visualizations appear in eFigures 4,5. Figure 3 presents qualitative comparisons for a representative PNOC patient with median cohort-level performance; Dice scores for each method are shown below the corresponding scans.

**Figure 2.**
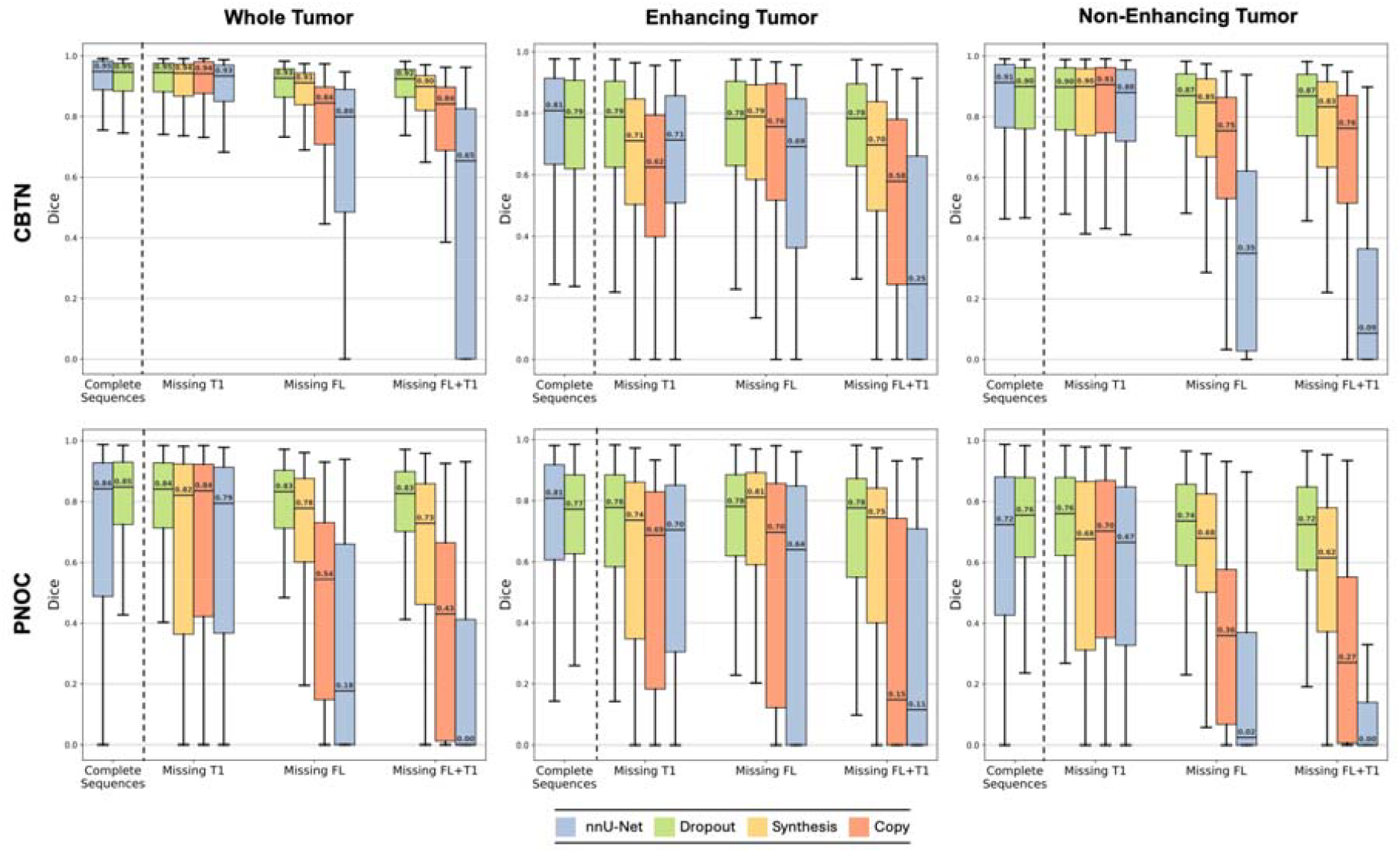
Segmentation Performance Under Complete and Missing MRI Scenarios Across Imputation Strategies: Box- and-whisker plots of Dice scores for WT, ET, and NET across imputation strategies and missing MRI scenarios in both the CBTN test set (top row) and PNOC clinical trial data (bottom row). Median Dice scores are annotated on each plot.

**Figure 3.**
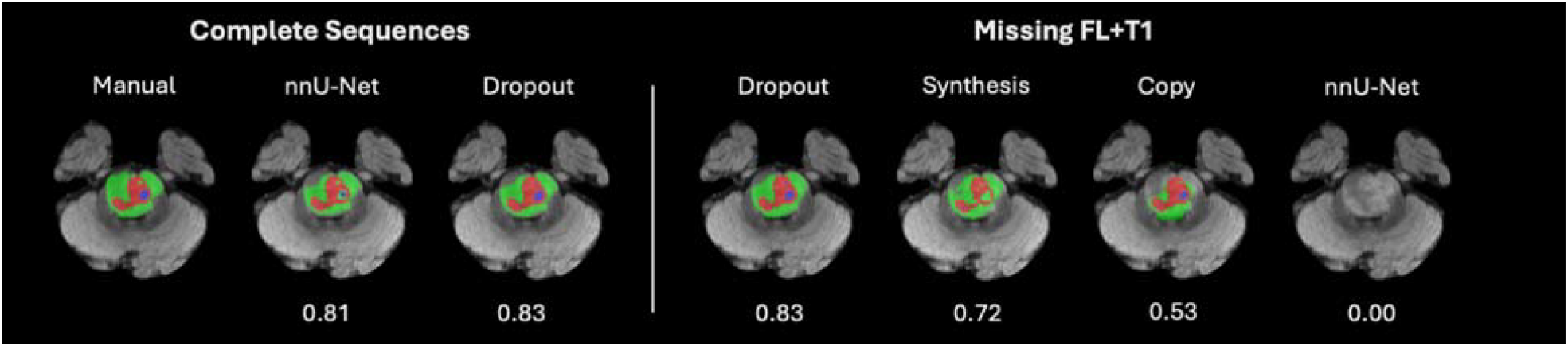
Representative Segmentation Results with Complete and Incomplete MRI Input: Segmentation outputs for a representative patient from the PNOC003 cohort under two scenarios: complete MRI input (left) and missing FLAIR and T1w-pre sequences (right). Segmentations are shown for each method, with Dice scores annotated below. Dice values reflect cases with performance closest to the cohort median in each setting. Tumor subregions are color-coded as follows: Green=NET, Red=ET, Blue=Cyst.

#### A. Complete MRI Sequences: Dropout Enhances Generalizability

On the CBTN test set (Figure 2), Dropout and baseline nnU-Net demonstrated comparable median/mean/std Dice scores (WT: 0.95/0.90/0.15 vs 0.95/0.90/0.13, NET: 0.91/0.82/0.22 vs 0.90/0.82/0.21, and ET: 0.81/0.73/0.24 vs 0.79/0.73/0.24) and interquartile ranges (IQRs). No significance was reached for ET (p adj. = 0.79, eTable 3) and although statistical significance (p adj.<0.05, eTable 3) was observed for WT and NET, the average per-patient Dice improvement for nnU-Net over Dropout was no more than 0.006 across the three regions (eTable 3), suggesting limited clinical relevance. These trends indicate that incorporating dropout during training does not degrade segmentation performance when complete input sequences are available. In fact, the use of dropout appeared to enhance generalizability reflected in increased Dice scores and reduced IQRs.

In more detail, in the PNOC cohort with complete sequences, dropout achieved notably smaller IQRs and improved median/mean/std Dice scores for WT (0.85/0.78/0.22 vs 0.84/0.69/0.31) and NET (0.76/0.71/0.23 vs 0.72/0.63/0.31) with statistical significance (p adj.=0.003 for WT and 0.006 for NET) and average per-patient Dice gain of approximately 0.1 over nnU-Net (eTable 3). For ET, though nnU-Net demonstrated higher median (0.80 vs 0.77), no statistical significance was observed (p adj.=0.18), with the two methods achieving equal means (0.70) and the Dropout method demonstrating a narrower IQR (0.26 vs 0.31) suggesting improved robustness (eTable 3). Altogether, these results suggest that training with modality dropout enhances model robustness and generalizability. Additional statistics can be found in (eTable 3).

#### B. Missing MRI Sequences: Dropout Yields Most Robust Performance

In most missing MRI scenarios (Figure 2), the Dropout model achieved the highest median Dice scores and narrowest IQRs. This advantage was especially evident in the most clinically relevant cases: ET segmentation with missing T1w-pre and NET segmentation with missing FLAIR. For ET, Dropout outperformed the next-best method (Synthesis) in both cohorts: median/mean/std of 0.79/0.72/0.24 vs 0.71/0.64/0.27 (CBTN), and 0.78/0.69/0.28 vs 0.74/0.60/0.33 (PNOC). For NET with missing FLAIR, Dropout achieved 0.87/0.79/0.23 vs 0.85/0.74/0.26 (CBTN) and 0.74/0.68/0.23 versus 0.68/0.61/0.27 (PNOC). All differences were statistically significant (p adj. < 0.05). Across all remaining regions and MRI availability scenarios Dropout achieved average per-patient Dice gains up to 0.12 over the next-best method and up to 0.57 over all others (eTable 4,5).

The Dropout model maintained segmentation performance under missing sequences comparable to its performance with complete inputs, both in median Dice and IQRs (Figure 2). Specifically, in key clinical cases: For ET with missing T1w-pre, the median/mean/std Dice scores for CBTN were 0.79/0.73/0.24 (complete) vs 0.79/0.72/0.24 (missing), and for PNOC, 0.77/0.70/0.27 vs 0.78/0.69/0.28. For NET under missing FLAIR, scores were 0.90/0.82/0.21 vs 0.87/0.79/0.23 in CBTN, and 0.76/0.71/0.23 vs 0.74/0.68/0.23 in PNOC. Although these differences were statistically significant, the average per-patient Dice drop never exceeded 0.03 and 0.04 when considering all remaining regions and MRI availability scenarios indicating minimal practical impact in segmentation performance (eTable 6,7).

The Dropout method was outperformed in median Dice only in two cases: Synthesis in ET segmentation with missing FLAIR (0.79 vs 0.78 for CBTN and 0.81 vs 0.78 for PNOC), and by Copy in NET with T1w-pre missing (0.91 vs 0.90 for CBTN). However, these differences were either not statistically significant or had average per-patient Dice difference ≤ 0.03 indicating no practical significance (eTable 4,5).

#### C. Synthesis Models Add Information Beyond Source Scans

In most missing MRI scenarios (Figure 2), the Synthesis approach outperformed the Copy method, achieving significantly (p adj.<0.05) higher median Dice and IQRs with average per-patient Dice gains of up to 0.26 (eTable 8,9). For WT and NET segmentation with T1w-pre missing Copy performed comparably or slightly better, though gains over Synthesis did not exceed 0.03 on average (eTable 8,9). These results suggest that the synthesis model generates anatomically meaningful information beyond what is present in nearest physical modality.

#### D. Sensitivity Analysis: Clinical Characteristics Affecting Segmentation Performance

To identify cases where missing T1w-pre or FLAIR sequences led to substantial segmentation degradation, we formed two targeted cohorts based on baseline nnU-Net performance: Patients with ≥ 0.1 Dice drop in ET when T1w-pre was missing, and in NET when FLAIR was missing. Radiologist review revealed distinct patterns: the cohort affected most by missing T1-pre was marked by patients with mildly enhancing tumors, diffuse edema, or hydrocephalus. The cases affected most by missing FLAIR sequence primarily tended to lack post-contrast enhancement on T1w-post. These patterns highlight clinical contexts where specific sequences are critical to accurate subregion delineation.

### Longitudinal Risk Stratification Using Clinical Trial Data

#### A. Automated Segmentation Under Complete Input Performs Prognostically Similar to Manual Segmentation

Figure 4, top shows Kaplan-Meier curves and corresponding forest plots based on tumor volumes from manual, baseline nnU-Net, and Dropout segmentations under complete input, showing progressively stronger separation between low/high-risk groups. The C-index modestly improves from 0.63 (manual), to 0.64 (baseline nnU-Net), to 0.65 (Dropout). Hazard ratios (HRs) also increased: 1.38 (p = 0.0541) for manual, 1.64 (p = 0.0045) for nnU-Net, and 1.68 (p = 0.0036) for Dropout. Risk scores did not differ significantly across models (p adj. > 0.05, eTable 10). The Dropout model yielded the most predictive combination, with tumor volume (HR = 1.68, p = 0.0036) and treatment group (HR = 2.57, p = 0.0076) emerging as significant covariates, indicating that both disease burden and therapeutic intervention play significant roles in determining patient’s survival.

**Figure 4.**
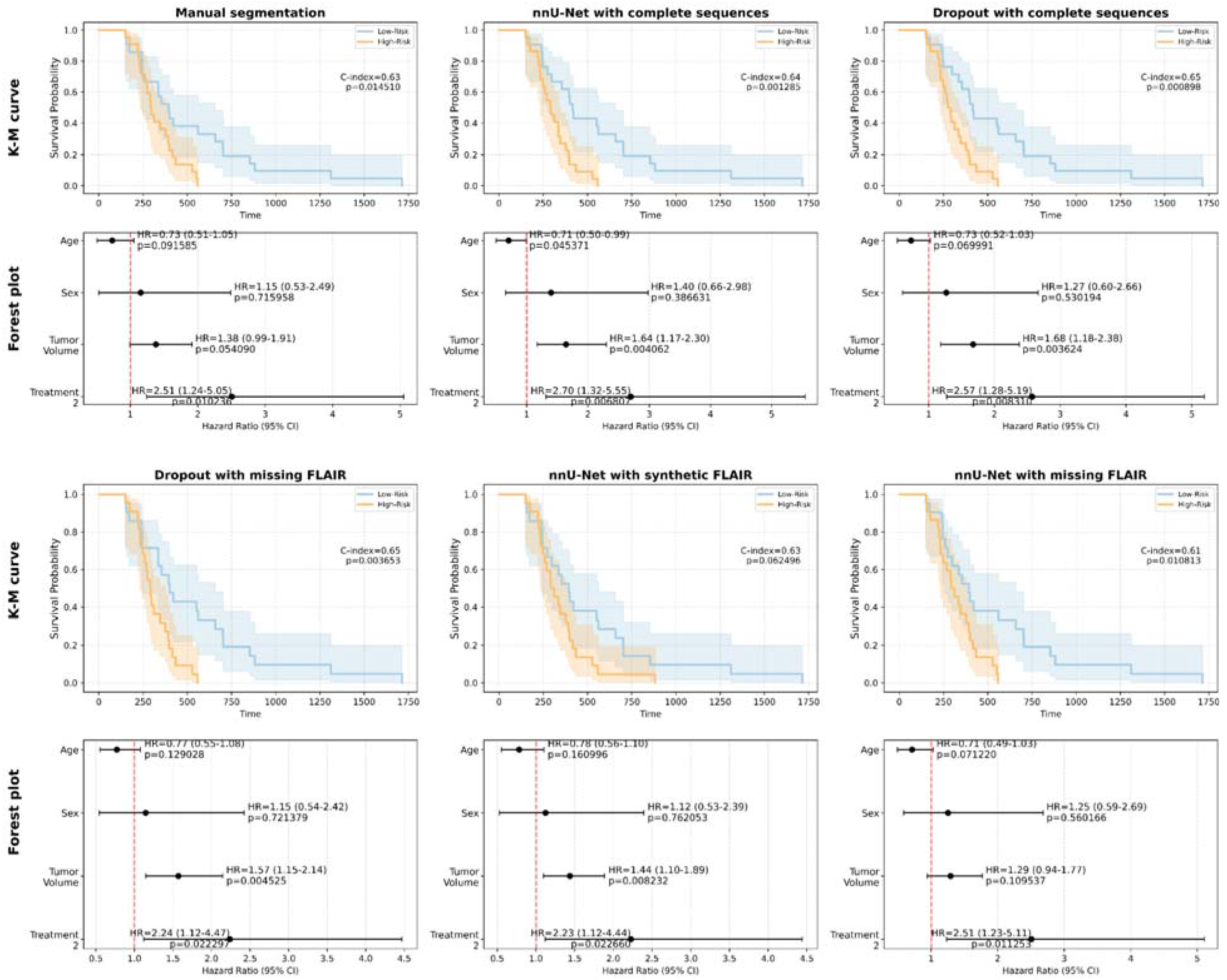
Prognostic Modeling Using Clinical Information and Tumor Volumes Derived from Manual and Automated Segmentations Under Complete and Incomplete MRI Inputs: *Top two rows*: Kaplan–Meier survival curves and correspondin forest plots from Cox models using age, sex, treatment group, and tumor volumes derived from manual segmentations, baseline nnU-Net, and Dropout with complete MRI input. *Bottom two rows: Corresponding Kaplan-Meier curves and forest plots under missing FLAIR, with segmentation handled via Dropout, Synthesis, or standard nnU-Net (missing inputs set to zero)*. Kaplan–Meier curves are annotated with the concordance index (C-index) and log-rank p-value. Forest plots show hazard ratios with 95% confidence intervals and p-values for each covariate.

#### B. Dropout with Missing FLAIR Remains the Most Robust Imputation Method

When FLAIR was missing (Figure 4, bottom), the Dropout model maintained a C-index of 0.65, and a significant association between tumor volume and survival (HR = 1.57, 95% CI: 1.15– 2.14, p = 0.0045), comparable to its performance with complete input. Risk scores from Dropout with missing FLAIR did not differ significantly from its complete-input counterparts (p > 0.05, eTable 10). In comparison, the Synthesis method achieved weaker results (C-index = 0.63; HR = 1.44, p = 0.0082), and baseline nnU-Net underperformed (C-index = 0.61; HR = 1.29, p = 0.106). Under missing FLAIR, risk scores from Dropout were statistically different (p adj.<0.05) from those produced by both the Synthesis and nnU-Net models. These findings demonstrate that dropout training provides a robust implicit imputation of missing FLAIR, outperforming both explicit synthesis and naïve omission. Age and sex were not significantly associated with survival across models while treatment group remained a statistically significant predictor.

### Perceptual Quality Analysis of Synthesized scans

The Synthesis model generated FLAIR and T1w-pre scans with high perceptual quality in both CBTN and PNOC cohorts. On CBTN, median SSI/MSE/PSNR were 0.93/0.003/27.1 in FLAIR synthesis and 0.91/ 0.015/20.4 for T1w-pre synthesis. On PNOC, median metrics were 0.91/0.004/24.9 for FLAIR synthesis and 0.90/0.014/20.9 for T1w-pre. Representative examples with perceptual metrics are shown in Figure 5, additional samples appear in eFigure 6.

**Figure 5.**
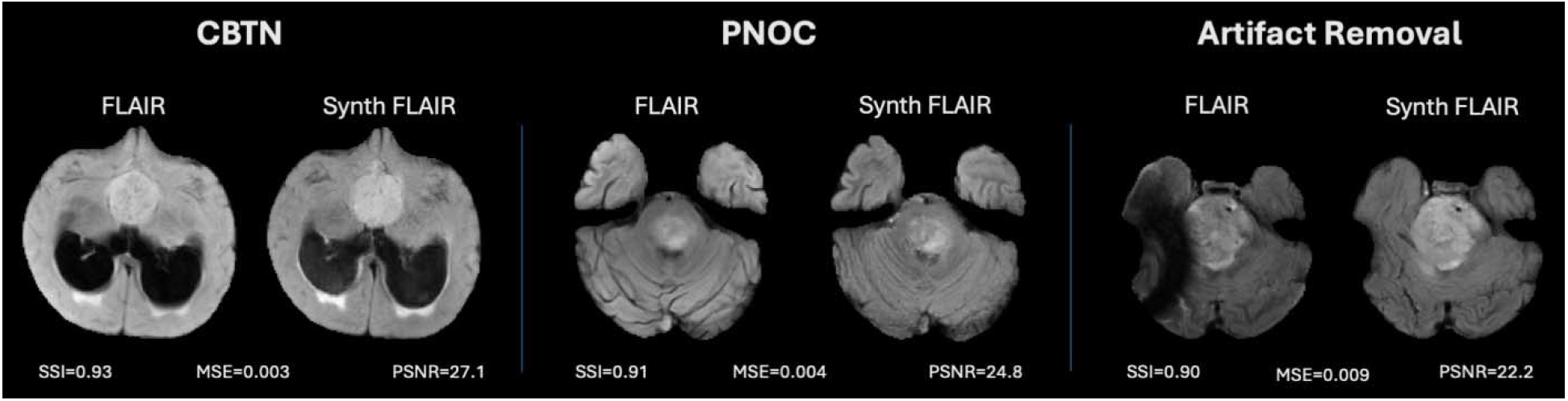
Representative Synthesis Examples from CBTN and PNOC: Shown are examples of synthesized FLAIR MRIs from the CBTN and PNOC cohorts, along with a case demonstrating artifact removal via synthesis. For each pair, the ground truth scan (left) and the corresponding synthesized scan (right) are shown, accompanied by their pairwise SSIM, MSE, and PSNR metrics.

In some cases, the synthetic images appeared visually superior to the ground truth, as the Synthesis method removed artifacts. Although these scans received lower perceptual scores due to dissimilarity with the artifact-containing ground truth, they exhibit higher visual quality. For instance, (Figure 5, right), a strong artifact in the ground truth FLAIR obscures part of the tumor, whereas the synthetic scan recovers its full extent.

## DISCUSSION

State-of-the-art DL segmentation models utilize the four standard MRI sequences (T1w-pre, T1w-post, T2w, and FLAIR), each providing unique information for tumor subregion delineation^11^. However, in clinical practice, MRIs exams are often incomplete due to acquisition variability or artifacts, a challenge amplified in pediatrics, where small cohort sizes are further reduced by missing data, hindering model generalizability.

Existing strategies such as MRI synthesis ^14^ or robustness-focused segmentation ^29^, are often unavailable to the public or validated only on adult datasets like BraTS ^30^, which are highly curated and not representative of pediatric clinical settings. To address these gaps, we leveraged a large, multi-institutional, multi-histology pediatric cohort of 715 patients from the CBTN^31,32^, BraTS-PEDs^8^ and externally validated on PNOC003/007 ^33,34^ clinical trials. We adapted the ResViT^14^ MRI synthesis model and developed a dropout-trained nnU-Net tumor segmentation model.

The dropout-trained model outperformed the out-of-the-box nnU-Net even under complete sequences, likely due to its more challenging training regime, where randomly dropped inputs forced the model to learn more robust and generalizable feature representations. In survival analysis, models built from Dropout-derived tumor volumes performed statistically comparable to those based on manual segmentations. Under missing sequences, the dropout-trained model consistently outperformed other imputation strategies, maintaining segmentation accuracy with minimal degradation, even when both T1w-pre and FLAIR were absent. Furthermore, under missing FLAIR, the model preserved prognostic risk stratification compared to complete MRI. This is particularly important for DMG/DIPG, which are predominantly non-enhancing tumors, making FLAIR imaging essential for accurate NET segmentation. DL-based MRI synthesis achieved high perceptual image quality, with anatomically consistent outputs, effective for artifact removal and qualitative assessment.

Despite these advances, this study has limitations. We selected ResViT for its strong performance in adult imaging studies, but its 2D architecture lacks volumetric context and requires extensive training and preprocessing, including skull-stripping and 2D slicing. While the dropout-based segmentation model demonstrated robustness to missing FLAIR and/or T1w-pre, its performance under additional missing MRI scenarios was not evaluated and warrants future investigation. Future work could explore more advanced generative architectures, such as 3D denoising diffusion probabilistic models (DDPMs), or many-to-one synthesis strategies that leverage multiple available sequences to reconstruct missing modalities.

In conclusion, we developed a robust modality imputation strategy using a dropout-trained segmentation model that performs reliably under both complete and incomplete MRI conditions. This approach offers a generalizable and practical solution for handling missing data in pediatric brain tumor segmentation and survival analysis, supporting more reliable AI deployment in real-world pediatric neuro-oncology settings.

## Data Availability

CBTN data and analysis results can be made available upon reasonable request

## REFERENCES

1. Jung AY. Basics for Pediatric Brain Tumor Imaging: Techniques and Protocol Recommendations. Brain Tumor Res Treat. 2024;12(1):1. doi:10.14791/btrt.2023.0037

2. Warren KE, Vezina G, Poussaint TY, et al. Response assessment in medulloblastoma and leptomeningeal seeding tumors: Recommendations from the Response Assessment in Pediatric Neuro-Oncology committee. Neuro Oncol. 2018;20(1):13–23. doi:10.1093/neuonc/nox087

3. Hoffman LM, Jaimes C, Mankad K, et al. Response assessment in pediatric craniopharyngioma: recommendations from the Response Assessment in Pediatric Neuro-Oncology (RAPNO) Working Group. Neuro Oncol. 2023;25(2):224–233. doi:10.1093/neuonc/noac221

4. Lindsay B, Besta NC, Milan I, et al. Response Assessment in Paediatric Intracranial Ependymoma: Recommendations from the Response Assessment in Pediatric Neuro-Oncology (RAPNO) Working Group. Vol 23.; 2022. https://www.thelancet.com/oncology

5. Poussaint TY, Mueller S, Franceschi E, et al. Title: Response Assessment in Pediatric High-Grade Glioma: Recommendations from the Response Assessment in Pediatric Neuro-Oncology Working Group.

6. Fangusaro J, Witt O, Hernáiz Driever P, et al. Response assessment in paediatric low-grade glioma: recommendations from the Response Assessment in Pediatric Neuro-Oncology (RAPNO) working group. Lancet Oncol. 2020;21(6):e305–e316. doi:10.1016/S1470-2045(20)30064-4

7. Cooney TM, Cohen KJ, Guimaraes C V, et al. Series Imaging Guidelines for Paediatric Brain Tumours 3 Response Assessment in Diffuse Intrinsic Pontine Glioma: Recommendations from the Response Assessment in Pediatric Neuro-Oncology (RAPNO) Working Group.; 2020. https://www.thelancet.com/oncology

8. Fathi Kazerooni A, Khalili N, Liu X, et al. BraTS-PEDs: Results of the Multi-Consortium International Pediatric Brain Tumor Segmentation Challenge 2023. Machine Learning for Biomedical Imaging. 2025;3(June 2025):72–87. doi:10.59275/j.melba.2025-f6fg

9. Fathi Kazerooni A, Arif S, Madhogarhia R, et al. Automated tumor segmentation and brain tissue extraction from multiparametric MRI of pediatric brain tumors: A multiinstitutional study. Neurooncol Adv. 2023;5(1):1–12. doi:10.1093/noajnl/vdad027

10. Familiar AM, Kazerooni AF, Vossough A, et al. Towards consistency in pediatric brain tumor measurements: Challenges, solutions, and the role of artificial intelligence-based segmentation. Neuro Oncol. 2024;26(9):1557–1571. doi:10.1093/neuonc/noae093

11. Vossough A, Khalili N, Familiar AM, et al. Training and Comparison of nnU-Net and DeepMedic Methods for Autosegmentation of Pediatric Brain Tumors. American Journal of Neuroradiology. 2024;45(8):1081–1089. doi:10.3174/ajnr.A8293

12. Madhogarhia R, Haldar D, Bagheri S, et al. Radiomics and radiogenomics in pediatric neuro-oncology: A review. Neurooncol Adv. 2022;4(1). doi:10.1093/noajnl/vdac083

13. Conte GM, Weston AD, Vogelsang DC, et al. Generative adversarial networks to synthesize missing T1 and FLAIR MRI sequences for use in a multisequence brain tumor segmentation model. Radiology. 2021;299(2):313–323. doi:10.1148/RADIOL.2021203786

14. Dalmaz O, Yurt M, Cukur T. ResViT: Residual Vision Transformers for Multimodal Medical Image Synthesis. IEEE Trans Med Imaging. 2022;41(10):2598–2614. doi:10.1109/TMI.2022.3167808

15. Cao B, Bi Z, Hu Q, et al. AutoEncoder-Driven Multimodal Collaborative Learning for Medical Image Synthesis. Int J Comput Vis. 2023;131(8):1995–2014. doi:10.1007/s11263-023-01791-0

16. Li Y, Shao HC, Liang X, et al. Zero-shot Medical Image Translation via Frequency-Guided Diffusion Models. Published online April 5, 2023. doi:10.1109/TMI.2023.3325703

17. Jiang L, Mao Y, Chen X, Wang X, Li C. CoLa-Diff: Conditional Latent Diffusion Model for Multi-Modal MRI Synthesis. Published online March 4, 2023. http://arxiv.org/abs/2303.14081

18. Özbey M, Dalmaz O, Dar SUH, et al. Unsupervised Medical Image Translation with Adversarial Diffusion Models. IEEE Trans Med Imaging. 2023;42(12):3524–3539. doi:10.1109/TMI.2023.3290149

19. Liu J, Pasumarthi S, Duffy B, Gong E, Datta K, Zaharchuk G. One Model to Synthesize Them All: Multi-Contrast Multi-Scale Transformer for Missing Data Imputation. In: IEEE Transactions on Medical Imaging. Vol 42. Institute of Electrical and Electronics Engineers Inc.; 2023:2577–2591. doi:10.1109/TMI.2023.3261707

20. Atli OF, Kabas B, Arslan F, et al. I2I-Mamba: Multi-modal medical image synthesis via selective state space modeling. Published online May 22, 2024. http://arxiv.org/abs/2405.14022

21. Li Y, Zhou T, He K, Zhou Y, Shen D. Multi-scale Transformer Network with Edge-aware Pre-training for Cross-Modality MR Image Synthesis. Published online December 2, 2022. doi:10.1109/TMI.2023.3288001

22. Arslan F, Kabas B, Dalmaz O, Ozbey M, Çukur T. Self-Consistent Recursive Diffusion Bridge for Medical Image Translation. Published online May 10, 2024. http://arxiv.org/abs/2405.06789

23. Chen C, Dou Q, Jin Y, Chen H, Qin J, Heng PA. Robust Multimodal Brain Tumor Segmentation via Feature Disentanglement and Gated Fusion. In: Shen D et al., ed. Medical Image Computing and Computer Assisted Intervention – MICCAI 2019. Springer, Cham; 2019:447–456. doi:10.1007/978-3-030-32248-9_50

24. Havaei M, Guizard N, Chapados N, Bengio Y. HeMIS: Hetero-Modal Image Segmentation. In: Medical Image Computing and Computer-Assisted Intervention – MICCAI 2016, Part II. Springer International Publishing; 2016:469–477. doi:10.1007/978-3-319-46723-8_55

25. Dorent R, Joutard S, Modat M, Ourselin S, Vercauteren T. Hetero-Modal Variational Encoder-Decoder for Joint Modality Completion and Segmentation. In: Medical Image Computing and Computer Assisted Intervention – MICCAI 2019. Springer, Cham; 2019:225–233. doi:10.1007/978-3-030-32245-8_9

26. Wang Y et al. ACN: Adversarial Co-training Network for Brain Tumor Segmentation with Missing Modalities. In: Medical Image Computing and Computer Assisted Intervention – MICCAI 2021. Springer, Cham; 2021:415–426. doi:10.1007/978-3-030-87234-2_39

27. Azad R, Khosravi N, Merhof D. SMU-Net: Style matching U-Net for brain tumor segmentation with missing modalities. In: International Conference on Medical Imaging with Deep Learning (MIDL 2022). Vol 172. PMLR; 2022:24–45. Accessed July 9, 2025. https://proceedings.mlr.press/v172/azad22a.html

28. Ding Y, Yu X, Yang Y. RFNet: Region-aware Fusion Network for Incomplete Multi-modal Brain Tumor Segmentation. In: Proceedings of the IEEE/CVF International Conference on Computer Vision (ICCV). IEEE; 2021:3975–3984.

29. Feng X, Ghimire K, Kim DD, et al. Brain Tumor Segmentation for Multi-Modal MRI with Missing Information. J Digit Imaging. 2023;36(5):2075–2087. doi:10.1007/s10278-023-00860-7

30. Baid U, Ghodasara S, Mohan S, et al. The RSNA-ASNR-MICCAI BraTS 2021 Benchmark on Brain Tumor Segmentation and Radiogenomic Classification. Published online July 5, 2021. http://arxiv.org/abs/2107.02314

31. Lilly J V., Rokita JL, Mason JL, et al. The children’s brain tumor network (CBTN) - Accelerating research in pediatric central nervous system tumors through collaboration and open science. Neoplasia (United States). 2023;35. doi:10.1016/j.neo.2022.100846

32. Familiar AM, Kazerooni AF, Anderson H, et al. A Multi-Institutional Pediatric Dataset of Clinical Radiology MRIs by the Children’s Brain Tumor Network.

33. Kline C, Jain P, Kilburn L, et al. Upfront Biology-Guided Therapy in Diffuse Intrinsic Pontine Glioma: Therapeutic, Molecular, and Biomarker Outcomes from PNOC003. Clinical Cancer Research. 2022;28(18):3965–3978. doi:10.1158/1078-0432.CCR-22-0803

34. Mueller S, Jain P, Liang WS, et al. A pilot precision medicine trial for children with diffuse intrinsic pontine glioma—PNOC003: A report from the Pacific Pediatric Neuro-Oncology Consortium. Int J Cancer. 2019;145(7):1889–1901. doi:10.1002/ijc.32258

35. Isensee F, Jaeger PF, Kohl SAA, Petersen J, Maier-Hein KH. nnU-Net: a self-configuring method for deep learning-based biomedical image segmentation. Nat Methods. 2021;18(2):203–211. doi:10.1038/s41592-020-01008-z

36. Lazow MA, Nievelstein MT, Lane A, et al. Volumetric endpoints in diffuse intrinsic pontine glioma: p correlations in the International DIPG/DMG Registry. Neuro Oncol. 2022;24(9):1598–1608. doi:10.1093/neuonc/noac037

37. Von Reppert M, Ramakrishnan D, Brüningk SC, et al. Comparison of volumetric and 2D-based response methods in the PNOC-001 pediatric low-grade glioma clinical trial. Neurooncol Adv. 2024;6(1). doi:10.1093/noajnl/vdad172

38. Goodfellow I, Bengio Y, Courville A. Deep Learning.

