## Supplemental Material for "AI-Powered Segmentation and Prognosis with Missing MRI in Pediatric Brain Tumors"

**TABLE OF CONTENTS**

1. DATA
A. Demographic Information for CBTN, BraTS-Peds, and PNOC003/007 cohort
B. Data pre-processing pipeline
2. TRAINING DETAILS
A. Modality Dropout: Training for Robustness to Missing Inputs
B. Image Synthesis: Generating Missing Sequences with ResViT
C. Zeroed Input: Standard nnU-Net Without Imputation
3. SEGMENTATION PERFORMANCE
A. Complete MRI Sequences: Dropout Enhances Generalizability
B. Missing MRI Sequences: Dropout Yields Most Robust Performance
C. Synthesis Models Add Information Beyond Source Scans
4. LONGITUDINAL RISK STRATIFICATION USING CLINICAL TRIAL DATA
5. PERCEPTUAL QUALITY ANALYSIS OF SYNTHESIZED SCANS

**1. DATA**

A. Demographic Information for CBTN, BraTS-PEDs, and PNOC003/007 cohorts

**eTable 1** Key Demographic information for the CBTN and BraTS-PEDs cohort

| **BraTS-PEDs data** | |
| --- | --- |
| Number of Patients | 403 |
| Histology | High-grade astrocytoma including DMG/DIPG |
| **CBTN data** | |
| Number of Patients | 312 |
| Age (mean/median/std) in years | 9.3/8.5/5.4 |
| Sex (Male/Female/Unknown) | 167(53%) /142(46%)/3(1%) |
| Histology | Number (% of cohort ) |
| Low-grade glioma/astrocytoma (WHO grade I/II) | 172 (55.13%) |
| Medulloblastoma | 90 (28.85%) |
| DMG/DIPG | 23 (7.37%) |
| Ependymoma, High-grade glioma/astrocytoma (WHO grade III/IV), Germinoma , Ganglioglioma, Craniopharungioma, Other, Meningioma, Sarcoma, Neurocytoma, Teratoma , Subependymal Giant Cell Astrocytoma (SEGA) | 27 (8.65%) |
| Institution | Number (% of cohort ) |
| The Children's Hospital of Philadelphia | 300 (96.15%) |
| Seattle Children's Hospital | 9 (2.88%) |
| University of Pittsburgh | 2 (0.64%) |
| Hackensack | 1 (0.32%) |

**eTable 2** Key Demographic information for the PNOC003 and PNOC007 cohorts

| **PNOC003 and PNOC007 data** | |
| --- | --- |
| Number of Patients | 43 |
| Number of Imaging Timepoints | 157 |
| Median number of imaging timepoints per patient | 3 |
| Median time interval in months between timepoints | 2.1 |
| Age (mean/median/std) in years | 8.79/8.0/4.11 |
| Sex (Male/Female) | 58%/42% |
| Histology | DMG/DIPG |
| Deceased/Censored | 43/0 |

B. Data Preprocessing

All images were co-registered and resampled to an isotropic resolution of 1 mm³ based on the anatomical SRI24 atlas, resulting in MRI volumes of size 240 x 240 x 155 using CaPTk software [1], [2]

**2. TRAINING DETAILS**

A. Modality Dropout: Training for Robustness to Missing Inputs

In the four scatter plots shown in the top two rows (zFL, zT1, zFL_zT1, and real) of eFigure 1, we display the median Dice score for WT segmentation on the validation set (N=85) as a function of the dropout probability (p = 0, 10, …, 100) applied independently to the T1w-pre and FLAIR sequences during training. For each plot, the corresponding MRI sequence(s) are set to zero during inference. For instance, in the zFL_zT1 condition, both FLAIR and T1w-pre are zeroed out for all 85 validation cases. When FLAIR is set to zero in the validation set, the model trained with p=0.9 (90%) achieves the highest median validation dice of 0.9111. Likewise, for missing T1w-pre, p=0.1 is optimal following the general trend of having all four sequences that show optimal dropout at p=0 and decreasing performance with increasing dropout. This is consistent with the fact that T1w-pre contributes less critically to whole tumor segmentation. When both FLAIR and T1 are missing, p=0.9 is optimal. To identify the optimal amount of dropout leading to robust performance in all MRI availability scenarios for each level of dropout, we take the average of the median dice in each of the four MRI availability scenarios. Using this plot, we identify p=0.4 as the optimal amount of dropout. p=0.4 strikes a balance between the increased levels of dropout needed for missing FLAIR and missing FLAIR and T1 (p=0.9) and the low levels of dropout needed under complete sequences (p=0) and missing T1 (p=0.1).

Despite these findings, it is important to note that the precise value of p is not critical for WT segmentation performance. For example, the average WT Dice across the four availability scenarios was 0.913 for p=0.4, while the lowest-performing non-zero dropout configuration (p=1.0) achieved a Dice of 0.9063—a marginal difference of just 0.007. However, the inclusion of dropout altogether is essential: the model trained with p=0 yielded an average Dice of 0.8048, representing a substantial drop of 0.11 compared to the p=0.4. This suggests that dropout serves as an effective regularizer, even if the precise value is less critical. Consequently, to reduce computational burden, future studies may consider training only at p=0.4 for WT segmentation.

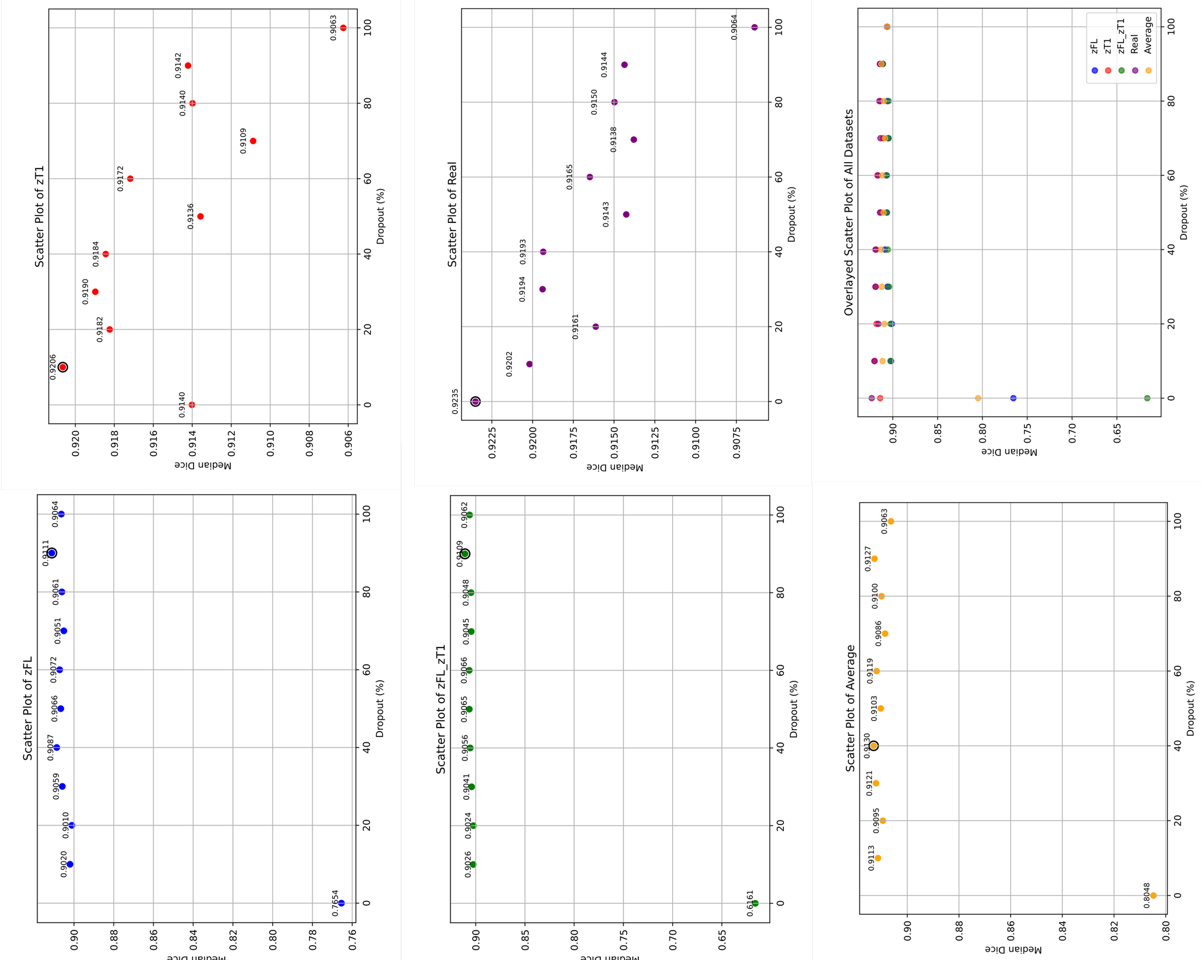

**eFigure 1**: Median WT Dice calculated in the validation set for various MRI availability scenarios as a function of dropout amount used during training.

B. Image Synthesis: Generating Missing Sequences with ResViT

In addition to standard image preprocessing (co-registered and resampled), each MRI volume was skull-stripped [3] and padded to a uniform shape of 256×256×155 using its background voxel intensity. Volumes were then normalized to the range [-1, 1]. To accommodate the 2D input format required by [4], each volume was sliced along the axial plane, resulting in 155 slices of size 1×256×256 per sequence. To train the synthesis models, T1w-pre slices were paired with their corresponding T1w-post slices, and FLAIR slices were paired with T2 slices. These paired datasets were used to train two separate ResViT models: one for synthesizing FLAIR from T2, and another for synthesizing T1w-pre from T1w-post.

Following the tuning protocol described in [4], we performed a grid search over learning rates in the set {$10^{-5}, 10^{-4}, 2\times10^{-4}, 5\times10^{-4}, 10^{-3}\}$ and loss weights in the set {20, 50, 100,150, 200, 250} to identify the parameters that maximixe PSNR in FLAIR from T2w synthesis in the validation set when trained for 100 epochs. The average PSNR value in the validation set as a function of epoch is shown in eFigure 2. The optimal hyperparameters for pediatric FLAIR MRI synthesis were found to be LR = $10^{-4}$ and PIX = 200. Notably, these values differ from the optimal settings reported for adult brain tumor synthesis in the BraTS dataset [5] (LR = $2\times10^{-4}$, weight = 100), underscoring the importance of domain-specific fine-tuning and reinforcing the observation that deep learning models trained on adult populations do not readily generalize to pediatric cohorts without careful adaptation. Following the ResViT protocol, the T1w-pre from T1w-post synthesis model used the same optimal hyperparameters determined in the FLAIR synthesis model (LR = $10^{-4}$ and PIX = 200).

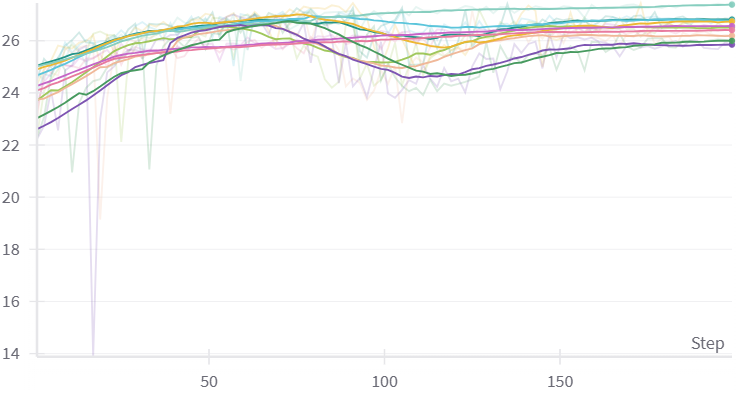

**eFigure 2**: Mean PSNR per epoch on the validation set for models taking hyperparameter values in the grid {$10^{-5}, 10^{-4}, 2\times10^{-4}, 5\times10^{-4}, 10^{-3}\}$ by {20, 50, 100,150, 200, 250}.

Following the recommendations of the authors of ResViT [4], we first pre-train the aggregated residual transformer (ART) blocks that synergistically combine residual convolutional (ResCNN) and transformer modules without the presence of Transformers to identify optimal hyperparameters (eFigure 2). We then fine-tune ResViT with the insertion of the Transformer modules, maintaining the hyperparameters determined through eFigure 2. As expected, the insertion of transformer modules lead to better performance compared to the ART blocks with only the ResCNN component. Hence, in eFigure 3 we that the “resvit” models (ART block with both transformer and ResCNN) give better mean PSNR per epoch than the “rescnn” (ART with only ResCNN) models.

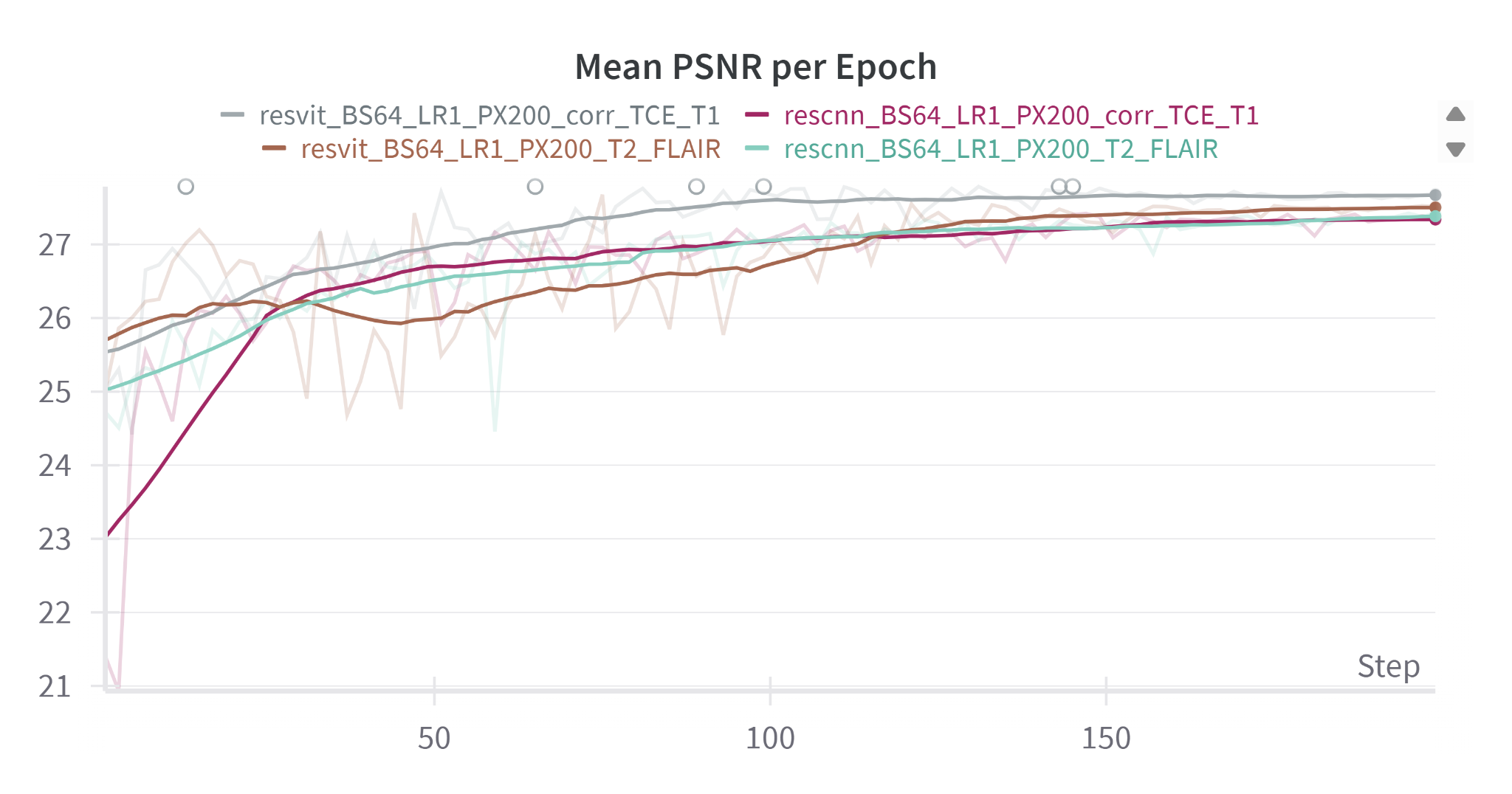

**eFigure 3:** Mean PSNR per epoch on the validation set for ResViT models with only pre-trained ART modules (“rescnn”) and ResViT models with the addition of transformer modules (“resvit”)

C. Zeroed Input: Standard nnU-Net Without Imputation

We trained a 3D full-resolution nnU-Net model using the default configuration of the open-source pipeline. Training was performed for 1000 epochs using stochastic gradient descent (SGD) with an initial learning rate of 0.01, momentum of 0.99, weight decay of 3e-5, and a polynomial learning rate scheduler. The loss function combined Dice and cross-entropy components, with deep supervision enabled throughout training. The self-configured architecture is a six-stage encoder-decoder U-Net architecture. Each encoder stage comprises two convolutional blocks, each consisting of a 3D convolution, instance normalization, and LeakyReLU activation. Feature channels increase across stages with widths of 32, 64, 128, 256, 320, and 320, and spatial resolution is reduced via strided convolutions. The decoder mirrors this structure with transposed convolutions for upsampling, skip connections from the encoder, and two convolutional blocks per stage. The final segmentation map is produced via a 1×1×1 convolution. The model was trained using patches of size 96×160×160 and Z-score normalized inputs across four MRI modalities. We take these values from debug.json and plans.json of the trained models.

**3. SEGMENTATION PERFORMANCE**

eFigures 4 and 5 show box-and-whisker plots of Dice scores for WT, ET, and NET across different imputation strategies and missing MRI scenarios in the CBTN and PNOC cohorts, respectively. Median Dice scores are annotated on each plot. The first row in each figure represents results for the entire cohort (N=290 for CBTN; N=167 for PNOC). The second row includes only patients whose manual segmentation masks contain the label of interest, for example, ET Dice scores are calculated only for patients with an ET label present in their manual mask, and likewise for NET. WT is by definition calculated on the entire cohort since all patients have at least one tumor subregion.This distinction provides a more accurate assessment of segmentation performance. Specifically, when evaluating ET and NET on the full cohort, Dice scores can be inflated (e.g. Dice=1.0) if the structure is absent in both the ground truth and the prediction, or deflated (e.g. Dice=0) when the model fails to detect a present structure. Providing both filtered and unfiltered results, clarifies whether the observed performance stems from the model's segmentation accuracy is influenced by the presence or absence of the target label. Nonetheless, overall performance trends remain consistent between the two representations. Figure 2 of the main text consists of the WT results along with label-present (second row) ET and NET values of eFigures 4 and 5.

Lastly, the PNOC cohort segmentation results contains a total of 167 timepoints, compared to the 157 reported in eTable 1. The demographics in eTable 1 reflect the subset used for survival analysis. While all 167 time points had valid segmentations suitable for Dice score evaluation, 10 were excluded from survival modeling due to temporal annotation inconsistencies that rendered them unsuitable for accurate longitudinal analysis.

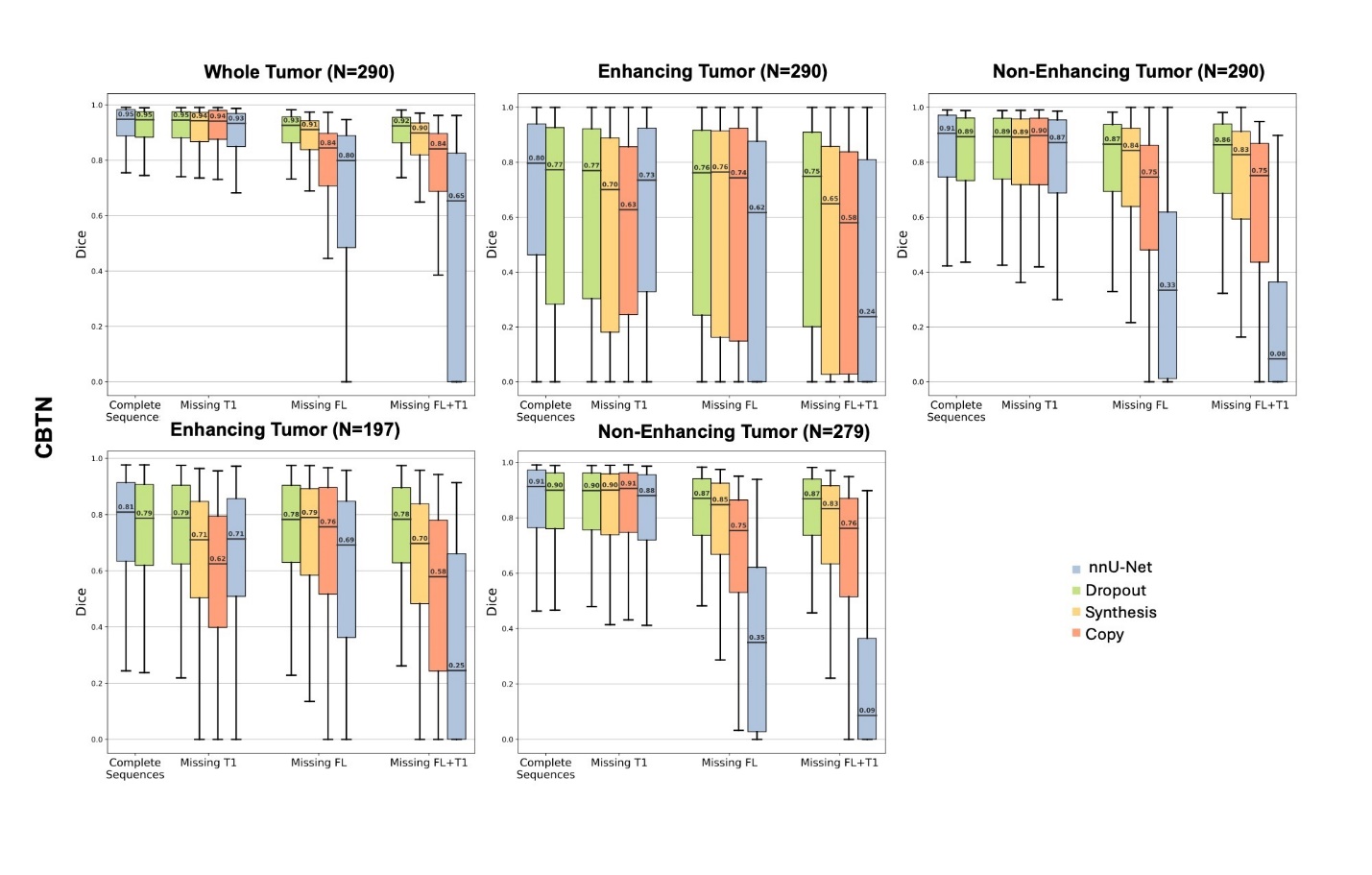

**eFigure 4:** Box-and-whisker plots of Dice scores for WT, ET, and NET across imputation strategies and missing MRI scenarios for the CBTN test set. Median Dice scores are annotated on each plot. Top row includes the entire cohort, second row includes only patients whose manual segmentation masks contain the label of interest.

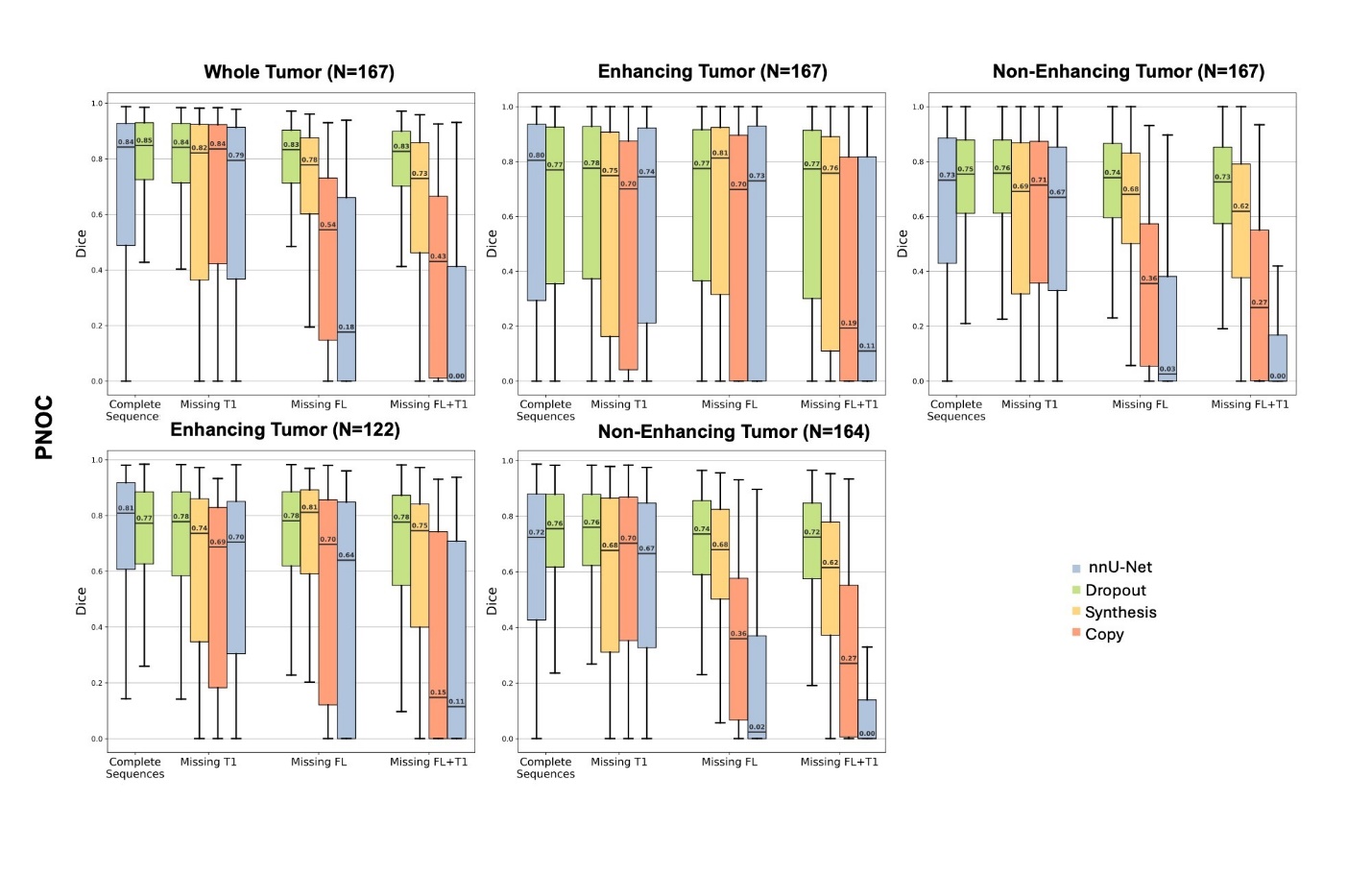

**eFigure 5:** Box-and-whisker plots of Dice scores for WT, ET, and NET across imputation strategies and missing MRI scenarios for the PNOC test set. Median Dice scores are annotated on each plot. Top row includes the entire cohort, second row includes only patients whose manual segmentation masks contain the label of interest.

The subsections below (A,B,C) provide supplemental statistics for Figure 2 of the main text.

A. Complete MRI Sequences: Dropout Enhances Generalizability

In eTable 3, we consider Dice statistics and significance tests between the performance of Dropout and out-of-the-box nnU-Net (Baseline) under complete sequences for WT, ET, and NET. We perform the Signed-Rank Wilcoxon test and display the Wilcoxon statistic (Wilcoxon_stat) and the associated raw p-value (Raw_p), and the adjusted p-value after Bonferroni correction (Bonferroni_p_adj). Additionally, we calculate the mean, median, standard deviation, and IQR for each of the two methods (Dropout, Baseline). “Wins” refer to the number of times one method achieved a higher Dice over the other method for a given patient. “Mean_Diff” is the average per-patient Dice difference of Method 1 minus Method 2. Positive values indicate higher Method 1 Dice scores. Lastly, we calculate the effect size r. r=0.10 implies small effect, 0.30 medium, 0.50+ large effect size.

**eTable 3** Dice Statistics and significance tests between the performance of Dropout and out-of-the-box nnU-Net (Baseline) under complete sequences for WT, ET, and NET.

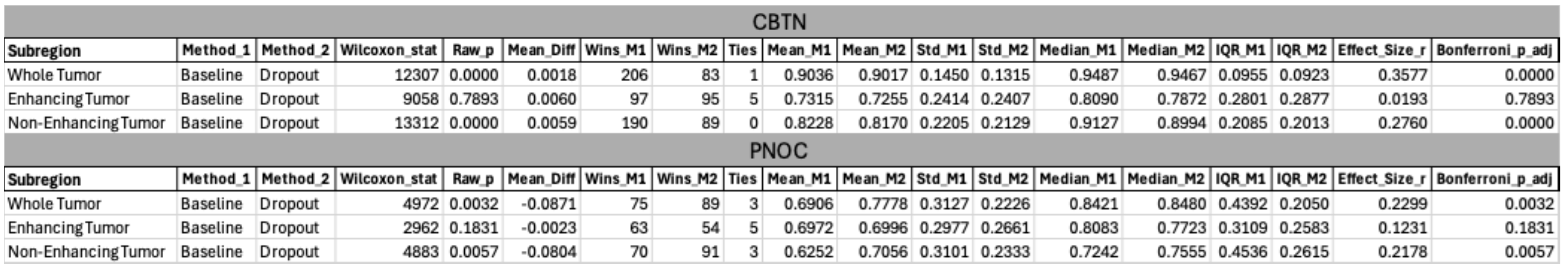

For **Whole Tumor (WT) subregion** on **CBTN**: nnU-Net and Dropout exhibit equal medians (0.95) and means (0.90), while the Dropout method achieves the smallest IQR (0.09 vs 0.1) and standard deviation (0.13 vs 0.15). Although the difference between the two methods is statistically significant (p<<0.05) with a moderate effect size (r = 0.36), the mean per-patient Dice improvement of only 0.002 in favor of nnU-Net is likely of minimal clinical relevance. Hence, for WT, the two methods perform on par.

For **Enhancing Tumor (ET) on CBTN,** nnU-Net achieves a higher median (0.81 vs 0.79) and lower IQR (0.28 vs 0.29), while the methods have equal averages (0.73) and standard deviations (0.24). There is no statistical significance (p =0.79), a negligible effect size (r = 0.02), and a small average per-patient Dice difference of 0.006 between the two methods. Hence, for ET, the two methods perform on par.

For **Non-Enhancing Tumor (NET) on CBTN,** nnU-Net achieves a higher median (0.91 vs 0.90) while both methods achieve the same mean (0.82). Dropout achieves lower IQR (0.20 vs 0.21) and standard deviation (0.21 vs 0.22). The difference reaches statistical significance with a small-to-moderate effect size (r = 0.28), yet the mean per-patient Dice difference remains clinically marginal at 0.006. For NET, the two methods perform on par.

For the **PNOC cohort**, for the **WT subregion**, the Dropout methods achieve a higher median value (0.85 vs 0.84), a higher mean (0.78 vs 0.70), a smaller IQR (0.21 vs 0.44), and a smaller standard deviation (0.22 vs 0.31). The difference between the two methods is statistically significant (p=0.003) with a small effect size (r= 0.23) and a mean per-patient Dice improvement of 0.08 (~0.1) in favor of Dropout. These trends support Dropout as the more effective method for WT segmentation on the PNOC cohort.

For the **ET subregion on PNOC**, the baseline method achieves a higher median (0.80 vs 0.77). Yet the two methods demonstrate the same mean (0.70), no statistical significance (p=0.18), a small effect size (r=0.12), and a marginal Dice improvement of 0.002 in favor of Dropout. Additionally, Dropout achieves lower IQR (0.26 vs 0.31) and lower standard deviation (0.27 vs 0.30). Here, we conclude that the methods perform on par for ET segmentation on the PNOC cohort, with increased robustness for the Dropout method due to smaller IQR and standard deviation.

For the N**ET subregion on PNOC**, the Dropout method achieves a higher median Dice (0.76 vs 0.72), higher average Dice (0.71 vs 0.63), lower IQR (0.26 vs 0.45), and lower standard deviation (0.23 vs 0.31). The difference is statistically significant (p=0.01) with a small effect size (r= 0.22) and a mean Dice improvement of 0.08 (~0.1) in favor of Dropout. These trends support Dropout as the more effective method for NET segmentation on the PNOC cohort.

B. Missing MRI Sequences: Dropout Yields Most Robust Performance

In eTables 4,5, we calculated the same statistics as in eTable 3. This time we compare the best (highest median) method vs the remaining methods on the CBTN and PNOC test sets for various missing MRI scenarios.

**eTable 4**: Best (highest median) method vs remaining methods on CBTN

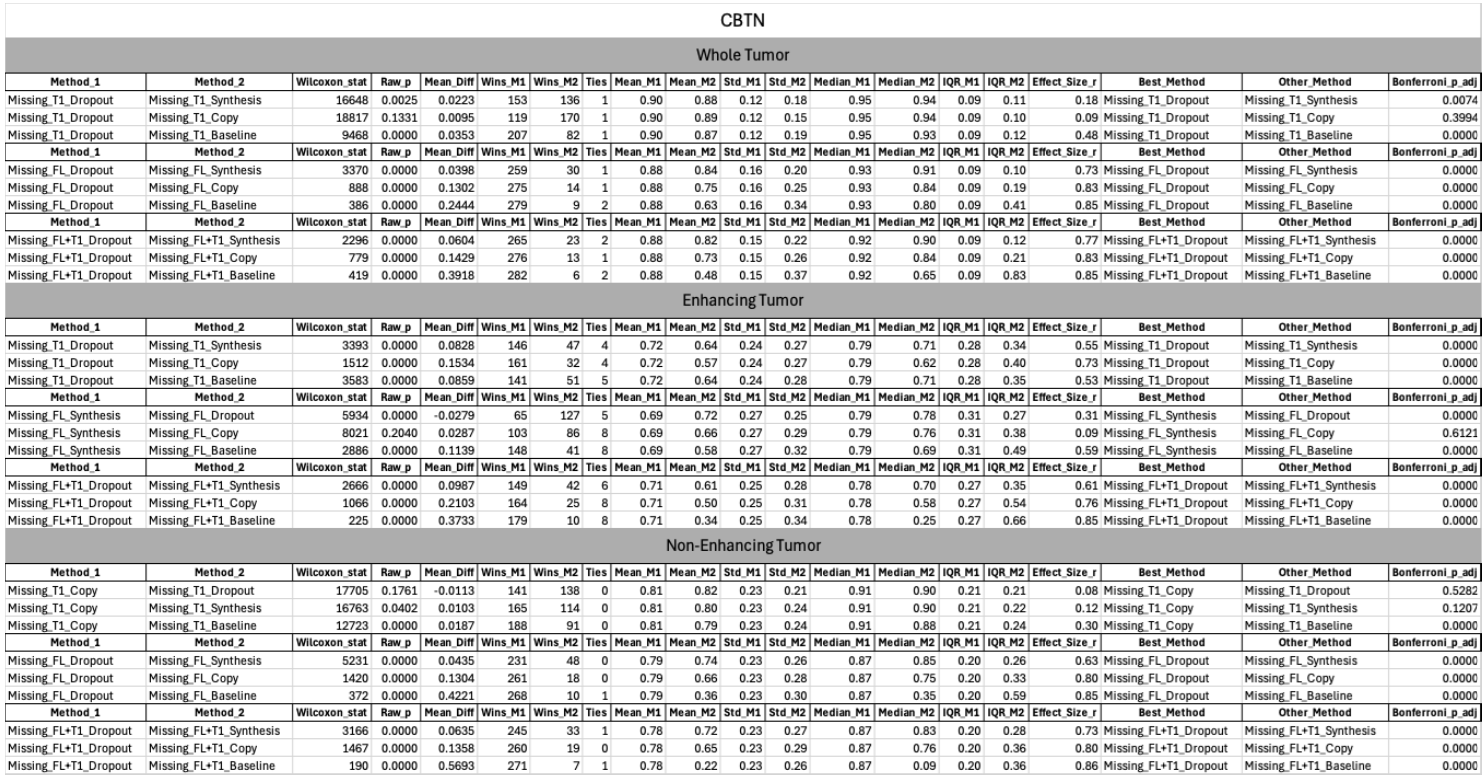

**eTable 5**: Best (highest median) method vs remaining methods on PNOC

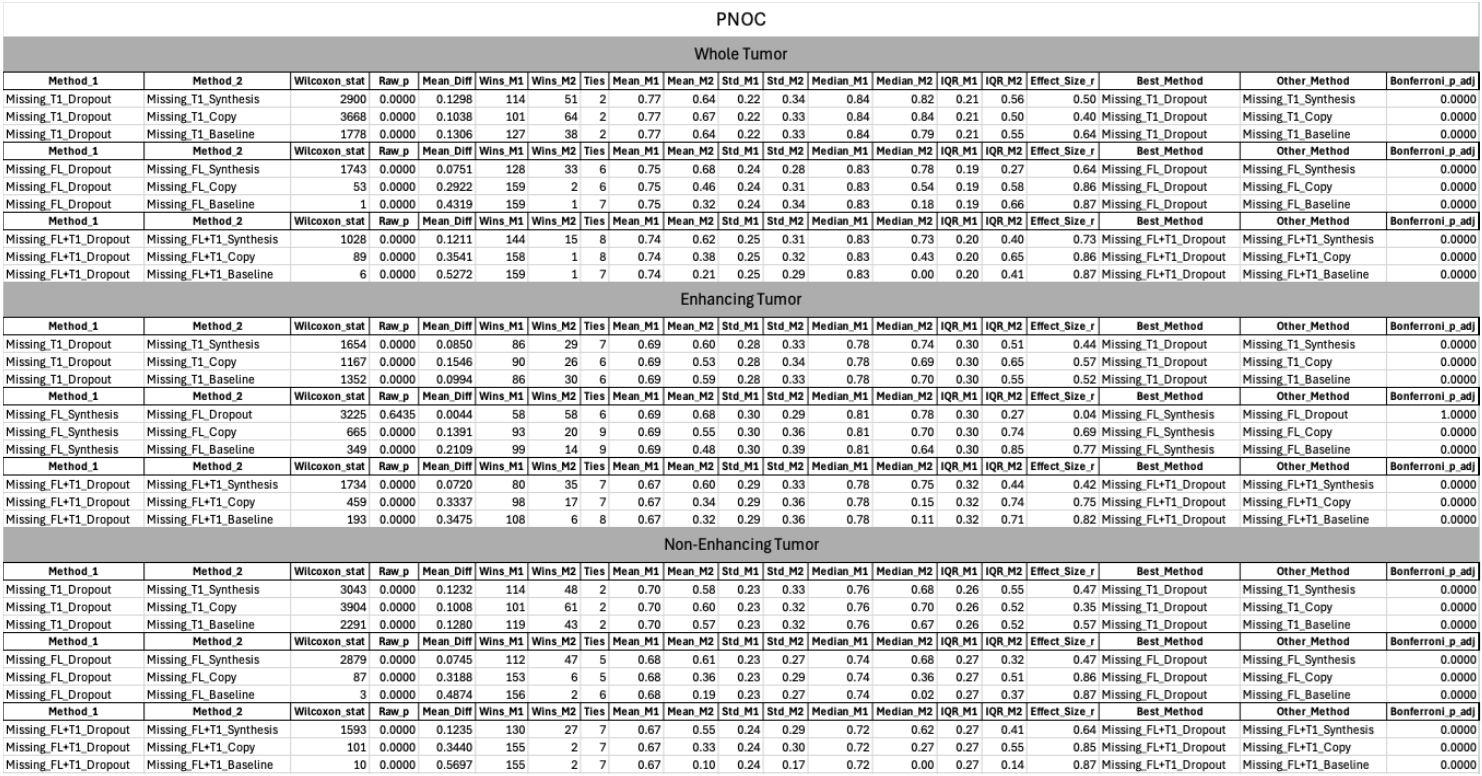

Comments for eTables 4, 5:

For the **CBTN** cohort, in **the WT** **subregion**, the Dropout method emerges as the most robust approach, consistently achieving the highest median Dice scores. When **T1** is missing, it yields marginal improvements over other methods, with mean per-patient Dice improvements ranging from 0.01 to 0.04. When **FLAIR** is missing, Dropout significantly outperforms alternatives (p << 0.05), showing large effect sizes ranging from 0.73 to 0.85, tighter IQRs, and mean per-patient Dice gains ranging from 0.04 to 0.24. When both **FLAIR and T1** are missing, again, Dropout statistically outperforms all alternative methods (p << 0.05) with tighter IQRs and effect sizes in the range 0.77-0.85 and mean per-patient Dice gains in 0.06-0.39.

For the **CBTN** cohort, in the **ET subregion,** when **T1** is missing, Dropout achieves the highest median Dice with strong statistical significance (p << 0.05), large effect sizes 0.53-0.73, and mean per-patient Dice gains 0.08-0.15. Likewise, when both **FLAIR and T1** are missing, Dropout achieves the highest median, large effect sizes 0.61-0.85, and mean per-patient Dice gains 0.1-0.37. When only **FLAIR** is missing, the Synthesis method reaches the highest median Dice score, yet Synthesis and Dropout practically perform equally well, with an average per-patient Dice increase of 0.03 (p < 0.05, r = 0.31) in favor of Dropout.

For the **CBTN** cohort, in the **NET** subregion, when **FLAIR** is missing, Dropout achieves the highest median Dice scores with strong statistical significance (p<<0.05), effect sizes of 0.63-0.85, and average per-patient Dice gains of 0.04-0.42 compared to alternatives. When both **FLAIR and T1** are missing, Dropout achieves the highest median Dice, large effect sizes of 0.73-0.86, and average per-patient Dice gains of 0.06-0.57. When **T1 is missing,** Copy leads in median Dice, but Dropout effectively performs equally with a mean per-patient Dice gain of 0.01 in favor of Dropout.

Conclusion: Dropout demonstrates the most reliable performance across all tumor subregions and missing MRI conditions in the CBTN cohort. In the majority of cases (except WT, missing T1, Synthesis), Dropout demonstrates statistical significance, and average per-patient Dice gains up to 0.1 compared to the second-best performing method.

In the **PNOC cohort**, in the **WT** subregion, Dropout achieves the highest median Dice scores for all missing scenarios, outperforming alternatives with statistical significance (p << 0.05). When **T1** is missing, Dropout achieves average per-patient Dice gains of 0.10-0.13 and large effect sizes of 0.40-0.64. When **FLAIR** is missing, Dropout achieves average per-patient Dice gains of 0.08-0.43 and large effect sizes of 0.64-0.87. When both **FLAIR and T1** are missing, Dropout achieves average per-patient Dice gains of 0.12-0.53 and large effect sizes of 0.73-0.87.

In the **PNOC cohort**, in the **ET** subregion, when **T1** is missing, Dropout achieves the highest median Dice scores with statistical significance (p<<0.05), average per-patient Dice gains of 0.09-0.15, and effect sizes of 0.44-0.57. When both **FLAIR and T1** are missing, Dropout achieves the highest median Dice scores with statistical significance (p<<0.05), average per-patient Dice gains of 0.07-0.35, and effect sizes of 0.42-0.82. When only **FLAIR** is missing, the Synthesis method produces the highest median Dice score, but the difference relative to Dropout is not statistically significant (p > 0.05) and yields a negligible average per-patient Dice gain of 0.004 in favor of Synthesis.

In the **PNOC** cohort, in the **NET** subregion, Dropout achieves the highest median Dice scores for all missing scenarios, outperforming alternatives with statistical significance (p << 0.05). When **T1** is missing, Dropout achieves average per-patient Dice gains of 0.12-0.13 and effect sizes of 0.35-0.57. When **FLAIR** is missing, Dropout achieves average per-patient Dice gains of 0.08-0.49 and effect sizes of 0.47-0.87. When both **FLAIR and T1** are missing, Dropout achieves average per-patient Dice gains of 0.12-0.57 and large effect sizes of 0.64-0.87.

In eTables 6,7, we calculated the same statistics as in eTable 3. This time we compare the Dice scores of the Dropout Method under Complete Sequences vs its performance under missing FLAIR, missing T1, and both FLAIR and T1 missing. In this way, we can evaluate how robust is the Dropout method in both complete and incomplete MRI sets.

**eTable 6**: Dropout Robustness across complete and incomplete MRI inputs on CBTN

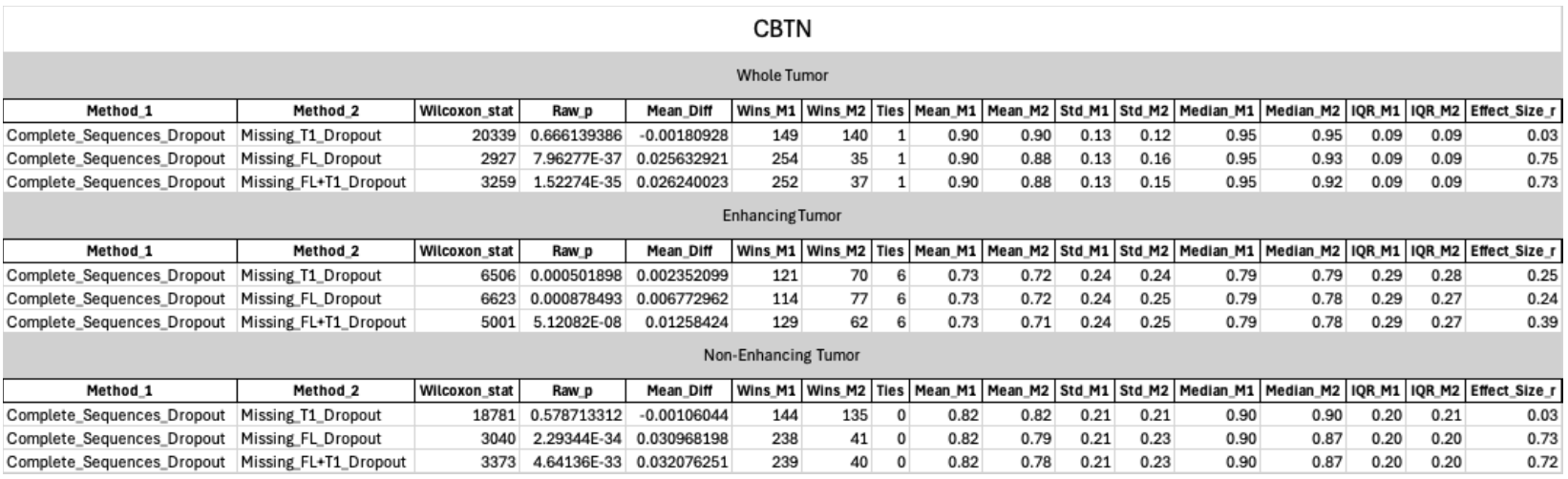

**eTable 7**: Dropout Robustness across complete and incomplete MRI inputs on PNOC

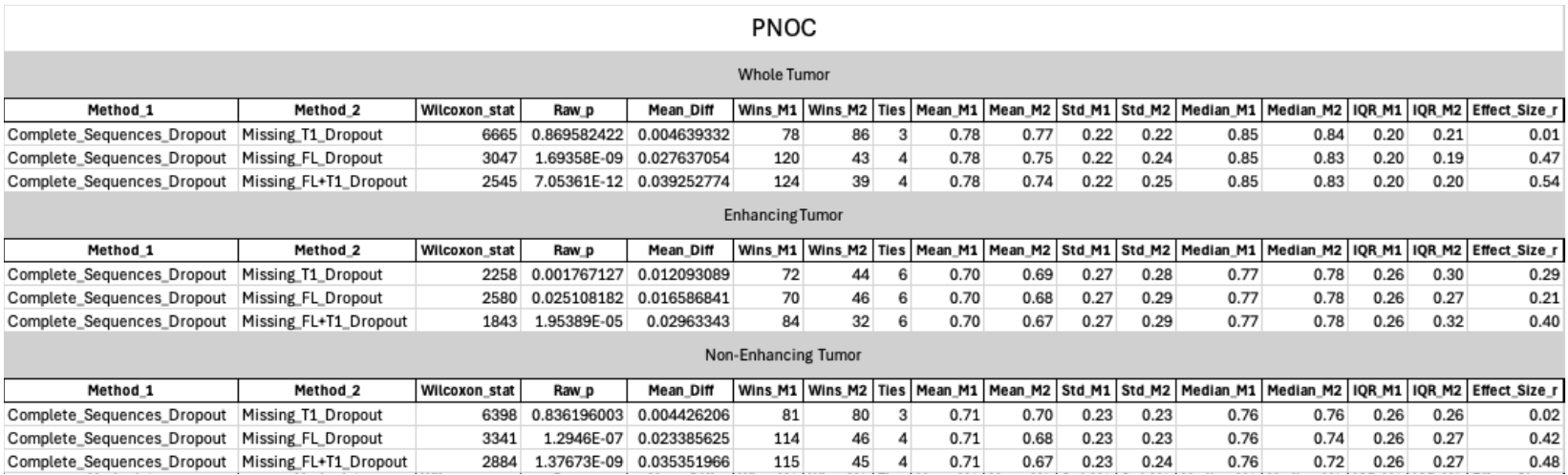

Comments for eTables 6, 7:

In the **CBTN** cohort, for the **WT** region, segmentation performance with **missing T1** is not statistically different from using the full set of MRIs (p = 0.67), with a negligible effect size (r = 0.03) and a clinically insignificant mean per-patient Dice difference of 0.002. When **FLAIR** is missing or both **FLAIR and T1** are missing, the Dice scores are statistically different (p<<0.05), yet the mean per-patient Dice differences are 0.03 for both cases, of negligible practical importance.

In the **CBTN** cohort for the **ET** subregion, when **T1, FLAIR, or both FLAIR and T1** are missing, segmentation performance with complete sequences vs incomplete sequences is statistically different (p<<0.05); however, again, we observe practically insignificant mean per-patient Dice differences ranging from 0.002 to 0.01.

In the **CBTN** cohort, for the **NET** subregion, segmentation performance with **missing T1** is not statistically different from using the full set of MRIs (p = 0.58), with a negligible effect size (r = 0.03) and a clinically insignificant mean per-patient Dice difference of 0.001. When **FLAIR** is missing or both **FLAIR and T1** are missing, the Dice scores are statistically different (p<<0.05), yet the mean per-patient Dice differences are 0.03 for both cases, again of negligible practical importance.

In the **PNOC** cohort, for the **WT** subregion, segmentation performance with **missing T1** is not statistically different from using the full set of MRIs (p = 0.87), with a negligible effect size (r = 0.01) and a clinically insignificant mean per-patient Dice difference of 0.005. When **FLAIR** is missing or both **FLAIR and T1** are missing, the Dice scores are statistically different (p<<0.05), yet the mean per-patient Dice differences are 0.03 and 0.04, respectively, again of negligible practical importance.

In the **PNOC** cohort for the **ET** subregion, when **T1, FLAIR, or both FLAIR and T1** are missing, segmentation performance with complete sequences vs incomplete sequences is statistically different (p<<0.05); however, again, we observe practically insignificant mean per-patient Dice differences ranging from 0.01 to 0.03.

In the **PNOC** cohort, for the **NET** subregion, segmentation performance with **missing T1** is not statistically different from using the full set of MRIs (p = 0.84), with a negligible effect size (r = 0.02) and a clinically insignificant mean per-patient Dice difference of 0.004. When **FLAIR** is missing or both **FLAIR and T1** are missing, the Dice scores are statistically different (p<<0.05), yet the mean per-patient Dice differences are 0.02 and 0.04, respectively, again of negligible practical importance.

C. Synthesis Models Add Information Beyond Source Scans

In eTables 8,9**,** we calculated the same statistics as in eTable 3. They are an expanded version of eTables 4,5. Instead of calculating comparisons between best methods vs the remaining, here we consider all possible pairs of methods for a given MRI availability scenario for CBTN and PNOC cohorts. We focus on Synthesis vs Copy methods in the main text.

**eTable 8** Pairwise Dice comparisons on CBTN

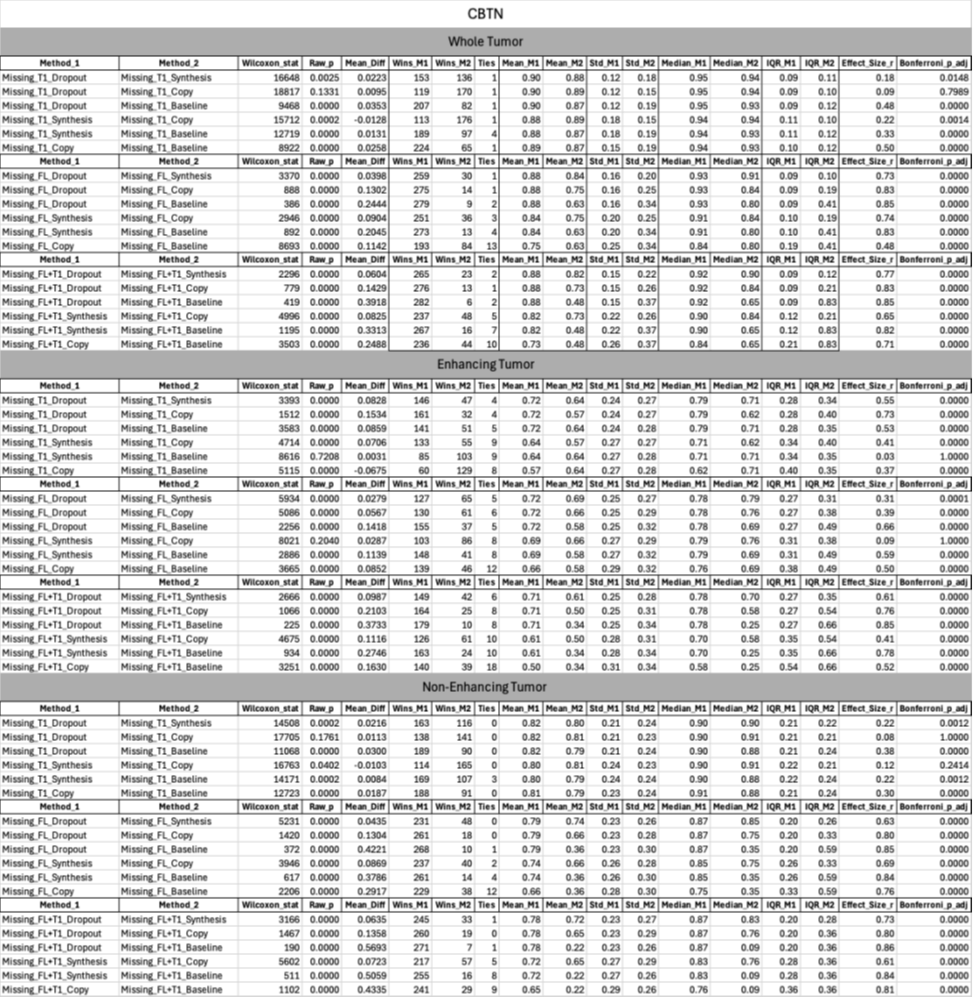

**eTable 9** Pairwise Dice comparisons on PNOC

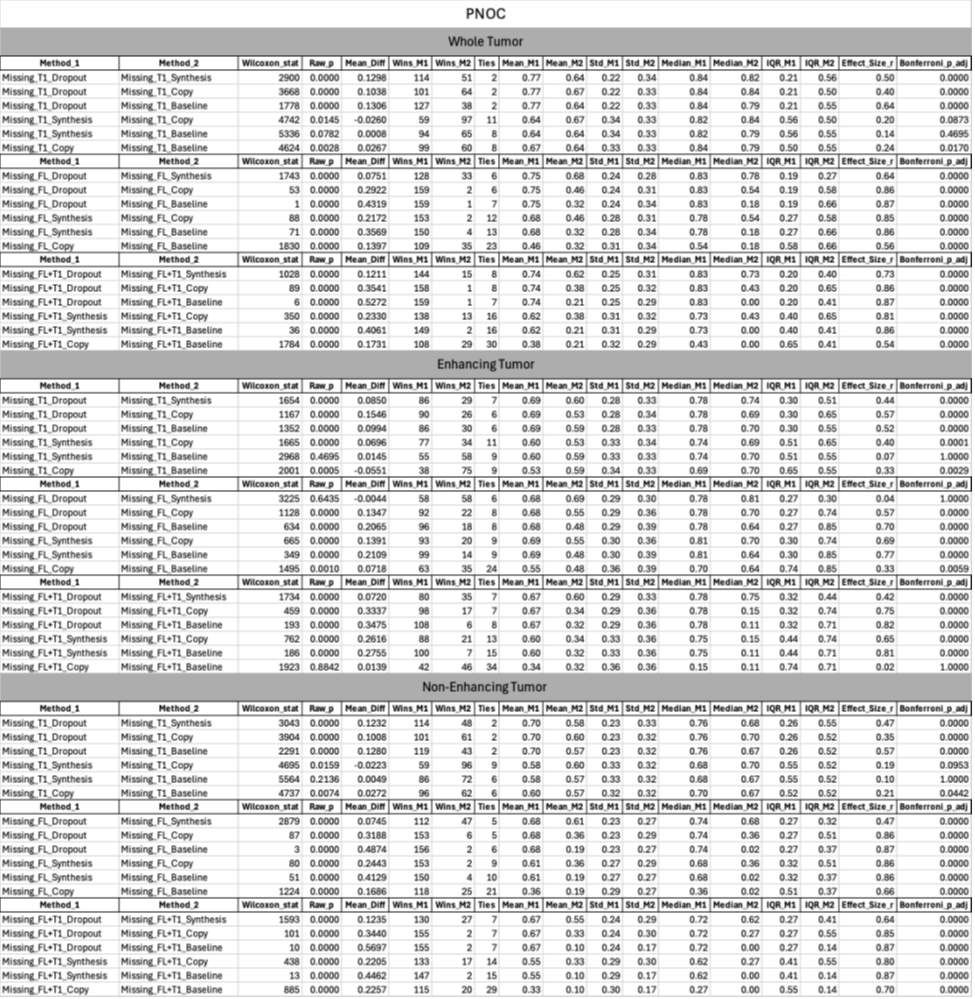

Comments on eTables 8,9 – Comparison of Synthesis vs Copy methods:

Notation: The first bracket at the end of a paragraph demonstrates the best method. In the second bracket, the first entry demonstrates the method with the highest median S=Synthesis, C=Copy, the second entry shows statistical significance (p) or its absence (-), and the last entry shows practical difference (pr) in average per-patient Dice gains. Dice changes near 0.1 are considerable.

On the **CBTN** cohort for **WT** segmentation:

For missing **T1,** both Synthesis and Copy achieve the same median (0.94); the distributions are statistically different, yet there is no practical difference with a mean per-patient Dice difference of 0.01. [Synthesis = Copy] [C/S,p,-]

For missing **FLAIR,** Synthesis achieves a higher median (0.91 vs 0.84)**.** The distributions are statistically different, with a mean per-patient Dice difference of 0.09 ~0.1 [Synthesis] [S,p,pr]

For missing **FLAIR and T1**, Synthesis achieves a higher median (0.90 vs 0.84). The distributions are statistically different, with a mean per-patient Dice difference of 0.08 ~0.1 [Synthesis] [S,p,pr]

On the **CBTN** cohort for **ET** segmentation:

For missing **T1,** Synthesis achieves a higher median (0.71 vs 0.62). The distributions are statistically different, with a mean per-patient Dice difference of 0.07 ~0.1 [Synthesis] [S,p,pr]

For missing **FLAIR,** Synthesis achieves a higher median (0.79 vs 0.76)**.** Yet there is no statistically significant difference, with a practically negligible mean per-patient Dice difference of 0.03 [Synthesis = Copy] [S,-,-]

For missing **FLAIR and T1** Synthesis achieves a higher median (0.70 vs 0.58). The distributions are statistically different, with a mean per-patient Dice difference of 0.11 [Synthesis] [S,p,pr]

On the **CBTN** cohort for **NET** segmentation:

For missing **T1,** Copy achieves a higher median (0.91 vs 0.90). The distributions are statistically different, yet there is no practical difference with a mean per-patient Dice difference of 0.01. [Synthesis = Copy] [C,p,-]

For missing **FLAIR,** Synthesis achieves a higher median (0.85 vs 0.75). The distributions are statistically different, with a mean per-patient Dice difference of 0.09 ~0.1 [Synthesis] [S,p,pr]

For missing **FLAIR and T1** Synthesis achieves a higher median (0.83 vs 0.76). The distributions are statistically different, with a mean per-patient Dice difference of 0.07 [Synthesis] [S,p,pr]

On the **PNOC** cohort for **WT** segmentation

For missing **T1,** Copy achieves a higher median (0.84 vs 0.82). The distributions are statistically different, yet there is no practical difference with a mean per-patient Dice difference of 0.03. [Synthesis = Copy] [C,p,-]

For missing **FLAIR,** Synthesis achieves a higher median (0.78 vs 0.54). The distributions are statistically different, with a mean per-patient Dice difference of 0.22 [Synthesis] [S,p,pr]

For missing **FLAIR and T1**, Synthesis achieves a higher median (0.73 vs 0.43). The distributions are statistically different, with a mean per-patient Dice difference of 0.23 [Synthesis] [S,p,pr]

On the **PNOC** cohort for **ET** segmentation

For missing **T1**, Synthesis achieves a higher median (0.74 vs 0.69). The distributions are statistically different, with a mean per-patient Dice difference of 0.07~0.1 [Synthesis] [S,p,pr]

For missing **FLAIR,** Synthesis achieves a higher median (0.81 vs 0.70). The distributions are statistically different, with a mean per-patient Dice difference of 0.14 [Synthesis] [S,p,pr]

For missing **FLAIR and T1** Synthesis achieves a higher median (0.75 vs 0.15). The distributions are statistically different, with a mean per-patient Dice difference of 0.26 [Synthesis] [S,p,pr]

On the **PNOC** cohort for **NET** segmentation

For missing **T1,** Copy achieves a higher median (0.70 vs 0.68). The distributions are statistically different, yet there is no practical difference with a mean per-patient Dice difference of 0.02. [Synthesis = Copy] [C,p,-]

For missing **FLAIR,** Synthesis achieves a higher median (0.68 vs 0.36). The distributions are statistically different, with a mean per-patient Dice difference of 0.24 [Synthesis] [S,p,pr]

For missing **FLAIR and T1,** Synthesis achieves a higher median (0.62 vs 0.27). The distributions are statistically different, with a mean per-patient Dice difference of 0.22 [Synthesis]. [S,p,pr]

**4. LONGITUDINAL RISK STRATIFICATION USING CLINICAL TRIAL DATA**.

This section presents supplemental statistics associated with Figure 4 of the main text. To assess whether the risk scores generated by different imputation strategies yield statistically meaningful differences, we employed non-parametric statistical tests. Because the risk scores were computed for the same cohort of patients across multiple methods, the data are paired and follow a repeated-measures design. Prior to comparison, we performed Shapiro-Wilk tests, which confirmed that the distributions of all risk scores significantly deviated from normality (p < 0.05). As a result, we used the Friedman test—a non-parametric alternative to repeated-measures ANOVA—to evaluate whether overall differences existed among the methods within each group. For pairwise comparisons between methods, we applied the Wilcoxon signed-rank test, which is suitable for matched, non-normally distributed data. To control for multiple hypothesis testing, we adjusted the resulting p-values using the Bonferroni correction. This approach provides a robust, assumption-free framework to determine whether different imputation strategies result in significantly different risk stratifications. In eTable 10, we consider three groups of comparisons associated with Figure 4 of the main text. In Group 1, we compare risk scores under complete MRI sets. In Group 2, we consider risk scores for Dropout, Synthesis, and baseline nnU-Net with missing FLAIR. In Group 3, we see how Dropout under complete sequences (that achieved the highest C-index) compares to Dropout, Synthesis, and baseline nnU-Net with missing FLAIR.

**eTable 10** Risk score statistics

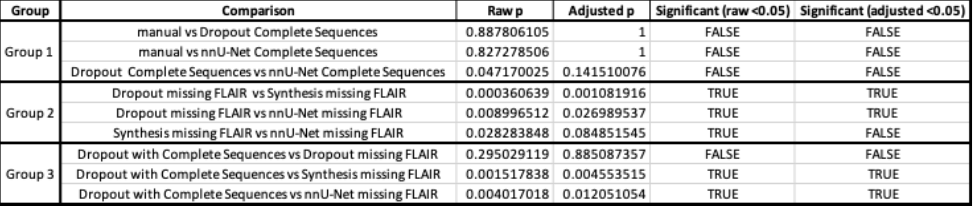

**5. PERCEPTUAL QUALITY ANALYSIS OF SYNTHESIZED SCANS**

eFigure 6 depicts cases of synthetic T1w-pre scans along with the FLAIR cases shown in the main manuscript. In the top right panel, the T1w-pre ground truth scan exhibits saturated intensities that obscure the boundary between gray matter, white matter, and the tumor, whereas the synthetic scan restores the contrast between these tissues. Additional visual inspection, reveal that the synthesized T1w-pre scans resemble, to some extent, the appearance of T1w-post scans, an expected outcome given the inherent challenge of removing contrast enhancement to recover the native T1w-pre appearance. This biophysically challenging task contributes to the lower perceptual metrics (e.g., MSE, PSNR, SSIM) observed for T1w-pre synthesis compared to FLAIR synthesis. Despite this, synthetic T1w-pre serves as a better substitute than simply copying the available T1w-post scan as shown in Segmentation Performance Section C of the main manuscript results. Hence, the synthesized T1w-pre scan function as a reliable substitution recovering information beyond what is present in the T1w-post input. This indicates the model’s capacity to synthesize features that are not merely interpolated but reflect realistic anatomical patterns.

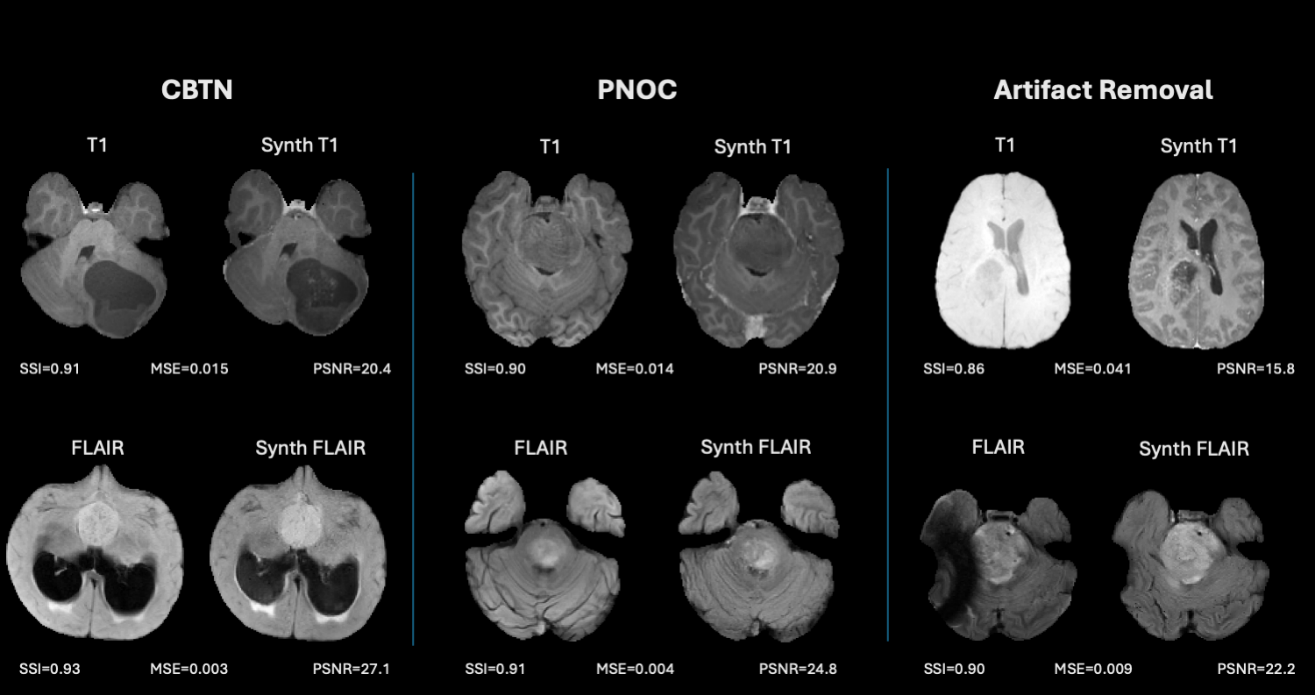

**eFigure 6:** Representative examples of synthesized T1 and FLAIR MRIs from the CBTN and PNOC cohorts, along with cases demonstrating artifact removal through synthesis. For each pair, the ground truth scan (left) and the corresponding synthesized scan (right) are shown, accompanied by their pairwise SSIM, MSE, and PSNR metrics.

**REFERENCES**

[1]	C. Davatzikos *et al.*, “Cancer imaging phenomics toolkit: quantitative imaging analytics for precision diagnostics and predictive modeling of clinical outcome,” *Journal of Medical Imaging*, vol. 5, no. 01, p. 1, Jan. 2018, doi: 10.1117/1.jmi.5.1.011018.

[2]	S. Pati *et al.*, “The cancer imaging phenomics toolkit (CaPTk): Technical overview,” in *Lecture Notes in Computer Science (including subseries Lecture Notes in Artificial Intelligence and Lecture Notes in Bioinformatics)*, Springer, 2020, pp. 380–394. doi: 10.1007/978-3-030-46643-5_38.

[3]	D. B. Gandhi *et al.*, “Automated pediatric brain tumor imaging assessment tool from CBTN: Enhancing suprasellar region inclusion and managing limited data with deep learning,” *Neurooncol Adv*, vol. 6, no. 1, Jan. 2024, doi: 10.1093/noajnl/vdae190.

[4]	O. Dalmaz, M. Yurt, and T. Cukur, “ResViT: Residual Vision Transformers for Multimodal Medical Image Synthesis,” *IEEE Trans Med Imaging*, vol. 41, no. 10, pp. 2598–2614, Oct. 2022, doi: 10.1109/TMI.2022.3167808.

[5]	U. Baid *et al.*, “The RSNA-ASNR-MICCAI BraTS 2021 Benchmark on Brain Tumor Segmentation and Radiogenomic Classification,” Jul. 2021, [Online]. Available: http://arxiv.org/abs/2107.02314
